# A computational model of CAR T-cell immunotherapy predicts leukemia patient responses at remission, resistance, and relapse

**DOI:** 10.1101/2021.09.21.21263913

**Authors:** Lunan Liu, Chao Ma, Zhuoyu Zhang, Weqiang Chen

## Abstract

Adaptive CD19-targeted CAR (Chimeric Antigen Receptor) T-cell transfer has become a promising treatment for leukemia. Though patient responses vary across different clinical trials, there currently lacks reliable early diagnostic methods to predict patient responses to those novel therapies. Recently, computational models achieve to *in silico* depict patient responses, with prediction application being limited. We herein established a computational model of CAR T-cell therapy to recapitulate key cellular mechanisms and dynamics during treatment based on a set of clinical data from different CAR T-cell trials, and revealed critical determinants related to patient responses at remission, resistance, and relapse. Furthermore, we performed a clinical trial simulation using virtual patient cohorts generated based on real clinical patient dataset. With input of early-stage CAR T-cell dynamics, our model successfully predicted late responses of various virtual patients compared to clinical observance. In conclusion, our patient-based computational immuno-oncology model may inform clinical treatment and management.

## Introduction

Anti-CD19 CAR (Chimeric Antigen Receptor) T-cell therapy is a promising immunotherapy for B cell acute lymphoblastic leukemia (B-ALL) (*1, 2*). Yet, clinical B-ALL cases demonstrated stochastically response and non-response to CAR T-cell therapy (*3*). For example, continuous/complete remission (CR) can be achieved in 70–90% of pediatric and adult patients, while long-term studies showed 30–60% of patients encounter either CD19-positive (CD19^+^) or CD19-negative (CD19^-^) relapse (*4*). Although various CAR T-cell products and combinational therapies have been tested in different clinical trials to improve patient response (*4, 5*), numerous questions remain unanswered to systematically understand the causes of the varied therapeutic responses. Thus, new clinical models that can predict patient responses to CAR T-cell treatment is critically desired to screen the most effective treatment protocol for individual patients.

Recently, computational models, based on empirical rationales, mathematic simulations with input of clinical data, provide valuable tools for *in silico* and systematical analysis of key biological mechanisms and patient responses in cancer immunotherapy (*6, 7*). Up-to-date, computational modeling of CAR T-cell therapy arises in early stages with applications of model-informed response prediction being limited. For example, a multiscale physiologically based pharmacokinetic-pharmacodynamic model had been developed for a quantitative study of relationship between CAR-affinity, antigen abundance, tumor cell depletion and CAR T-cell expansion using data collected from xenograft mouse models (*8*). Other approaches focus mostly on modeling factors underlying CAR T-cell dynamics, such as ecological dynamics regulated CAR T-cell explain expansion (*9*) and exhaustion (*10*), signaling-induced cell state variability (*11*), and CAR T-cell expansion due to lymphodepletion and competitive growth between CAR T-cell and normal T-cell (*12*). Lately, Liu et al. developed a model to characterize clinical CAR T-cell kinetics across response status, patient populations, and tumor types, yet only in a retrospective manner (*13*). However, these computational models typically fail to provide a collective analysis and effective interpretation of clinical trial data from different clinical studies to reveal the key cellular mechanisms underlying the heterogeneous patient outcomes observed in different clinical trials. Critically, a clinical data-based prognostic model that can predict patient responses to CAR T-cell treatment at early stage is largely absent.

In the present study, we developed a mathematical framework of CAR T-cell therapy structured with a matrix of numerical differential equations for a quantitatively study and *in silico* modeling of key biological mechanisms in CAR T-cell therapy, such as leukemia cell growth and apoptosis, CAR T-cell activation, expansion, and cytotoxic efficiency, and CD19 antigen mediated relapse mechanisms. After calibration and validation with clinical data from 209 leukemia patients, our computational model revealed key determinants that depicted the heterogeneous clinical responses between the responders, non-responders, and patients with CD19^+^/CD19^-^ relapse. A useful model that can provides accurate predictions requires large representative datasets for validation. Clinical trial simulation studies the effects of a therapy in virtual patient cohorts using mathematical models of physiological system, which to some extent enlarges sampling for clinical trials and full range of mechanistic testing (*14, 15*). Incorporating this concept, we performed a clinical trial simulation of CAR T-cell therapy using virtual patient cohorts generated based on real clinical patient dataset. With input of early-stage clinical and *in silico* generated virtual patient data, our model successfully predicted the late therapeutic outcomes of most patients under CAR T-cell treatment, which may inform clinical treatment and guide clinical trials of personalized CAR T-cell therapy.

## Results

### A computational model of CAR T-cell immunotherapy

To construct a computational model of CAR T-cell therapy that reproduces the pathophysiological processes and immunological interactions, we framed a matrix of numerical differential equations (Details see ‘Model construction’ in **Methods**) and calibrated the model using the stochastic approximation expectation maximization (SAEM) algorithm for nonlinear mixed-effect modeling (NLME), as implemented in Monolix (Lixoft, France). To fit and calibrate our model, we searched and collected clinical data of 209 B-ALL patients from 10 clinical trials of anti-CD19 CAR T-cell therapies and sampled into 32 groups (Details see ‘Collecting and processing clinical data’ in **Methods**). Of note, only statistic values like medians were available for several trials (**Supplemental Table S1-4**). The clinical data were assigned into different patient cohorts, i.e., CR, non-response (NR), CD19^+^ relapse and CD19^-^ relapse (**Figure 1**), based on the flow cytometry and quantitative polymerase chain reaction (qPCR) monitoring of CAR T-cell in the blood, as well as morphological testing of leukemia burden in the bone marrow. To minimize the variation across different batches and clinical trials, we unified and transformed all the clinical data at a scale equivalent to the bone marrow level (Details see ‘Collecting and processing of clinical data’ in **Methods**). Model fitting and parameterization processes is provided accordingly (Details see ‘Parameter estimation’ in **Methods**). For a dataset including different individuals, SAEM firstly fits the data as a group and estimate parameters on population level (population level parameters). After that, random effect is adopted to estimate the parameters of each individual (individual level parameters) based on population level parameters.

**Figure 1.**
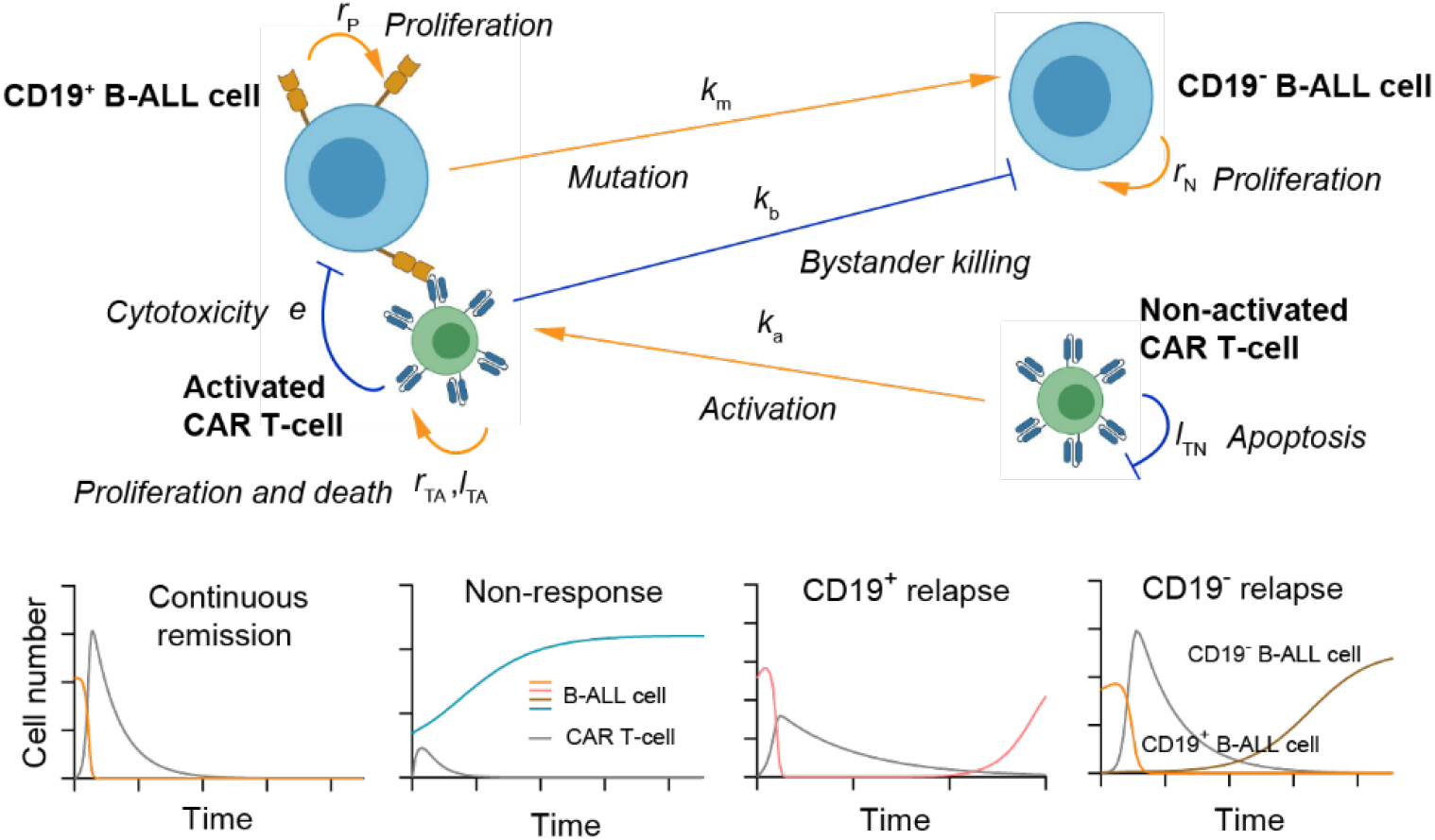
A schematic showing the key cellular components and their dynamic interactions in the computational CAR T-cell therapy model. Based on the computational model, four kinds of responses (bottom panel): continuous remission, non-response, CD19^+^ relapse and CD19^-^ relapse can be recapitulated with the outputs of dynamics of CAR T-cell and B-ALL cell.

We firstly calibrated the proposed computational model with clinical data of CAR T-cell and tumor burden of 148 CR patients (**Supplemental Table S1, Supplemental Figure S1**), and simulated the dynamic behaviors of CAR T-cell and tumor cell during the treatment. After being activated by the CD19^+^ B-ALL cells, CAR T-cell rapidly expanded and reached the peak within the first 1-2 weeks, and then gradually decreased after tumor cells were rapidly depleted. The results show a great correlation between clinical statistics and simulation data (**Figure 2A&B**). We found that the peak value of CAR T-cell during treatment increased as initial tumor burden increased, in consistent with clinical observation (**Figure 2C**). This is largely due to the rapid *in vivo* CAR T-cell expansion stimulated by CD19^+^ tumor cell as verified by the real-time simulation results with increasing initial tumor burden (**Figure 2D**). Similar correlation was seen between the peak value and activation rate of CAR T-cell (**Figure 2E**). Moreover, the day when CAR T-cell peaked is correlated to the day when patient achieved minimal residual disease (MRD^-^, tumor burden <0.01%) (**Figure 2F**).

**Figure 2.**
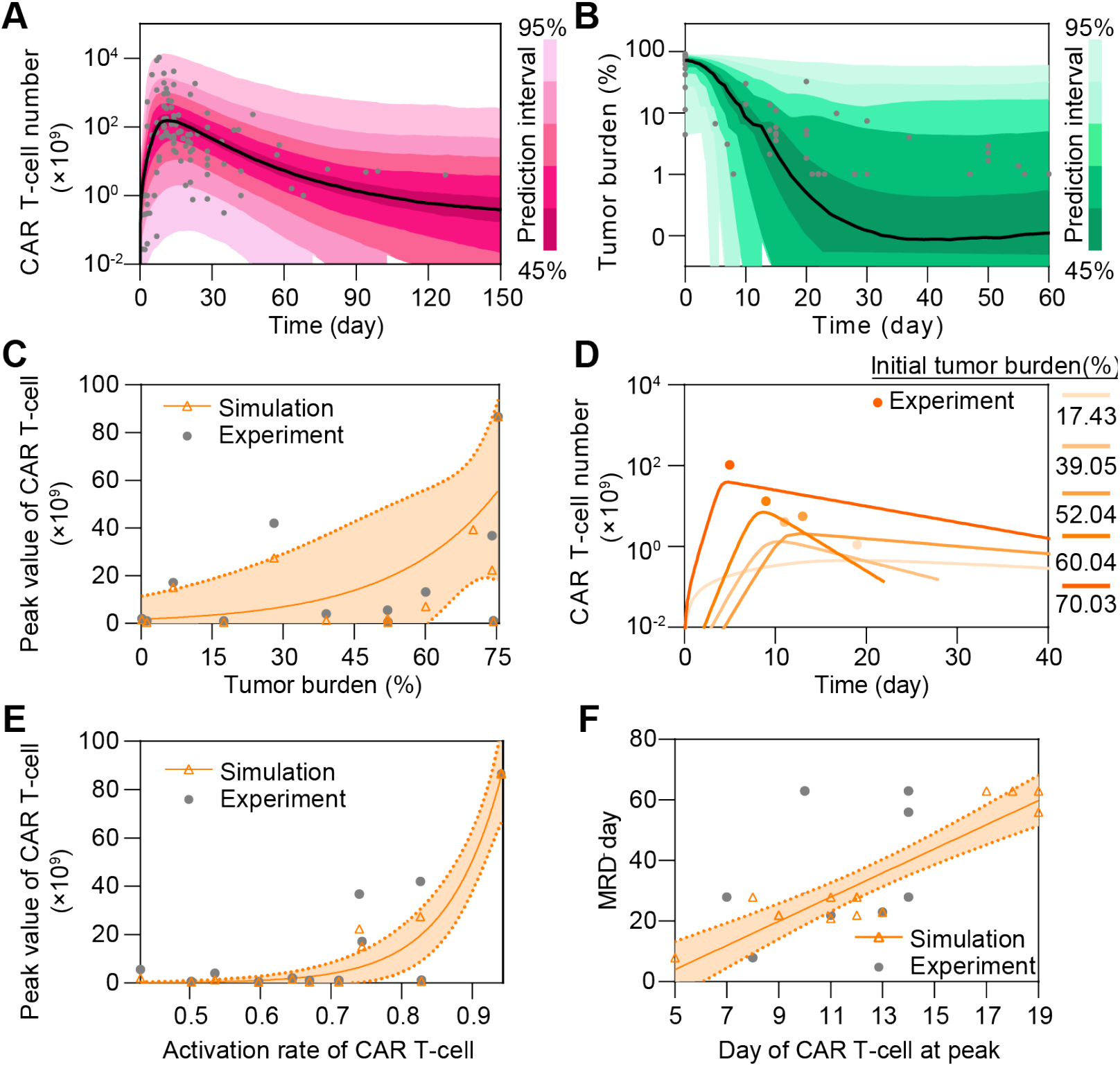
*In silico* analysis of continuous remission to CAR T-cell therapy. (**A&B**) Calibration results of CAR T-cell (**A**) and tumor burden (**B**) of remission patients with median (solid line) and 95% prediction interval (color bands). The dots represent the experimental data. Prediction interval was automatically generated by the predefined algorithm of Monolix software during calibration. (**C**) Relationship between tumor burden and peak value of CAR T-cell and (**D**) corresponding real-time results. Simulation results of different patient groups were calculated and fitted with dashed lines, and compared with experimental results in D. The band in C represents the 95% confidence interval (the same below). (**E&F**) Relationships between the activation rate and peak value of CAR T-cell (**E**), and day of CAR T-cell at peak and MRD^-^ day (**F**).

In parallel, we calibrated the model with clinical data of 24 NR patients to model the scenario of non-responders, which again confirmed its validity (**Figure 3A&B, Supplemental Figure S2, Supplemental Table S2**). For instance, significant differences were reproduced as observed in CR and NR patients in terms of CAR T-cell and tumor cell dynamics (**Figure 3C**). In particular, the peak value and AUC28 (area under the curve from day 0 to day 28, a commonly-used clinical marker to evaluate the expansion and function of CAR T) of CAR T-cells in NR patients were less than those of CAR T-cells in CR patients at population level. Similar trends were further confirmed at individual level by experimental and simulation data (**Figure 3D&E**). To understand such difference, we further ran a sensitivity analysis of population level parameters and found that those related to CAR T-cell functionality, such as growth rate *r*_TA_, killing rate *e*, activation rate *k*_A_, and apoptosis rate *l*_TA_, critically influence CAR T-cell therapy outcomes (**Supplemental Figure S3**). Our model indicated that the median values of these factors significantly differed between CR and NR patients (**Figure 3F-I**). CAR T-cell in CR patients but not those in NR patients generally have higher growth rate *r*_TA_, killing rate *e*, activation rate *k*_A_ but lower apoptosis rate *l*_TA_, suggesting an impaired CAR T-cell functionality in NR patients. These results together demonstrate the capability of our computational model to recapitulate the clinical dynamics during CAR T-cell therapy.

**Figure 3.**
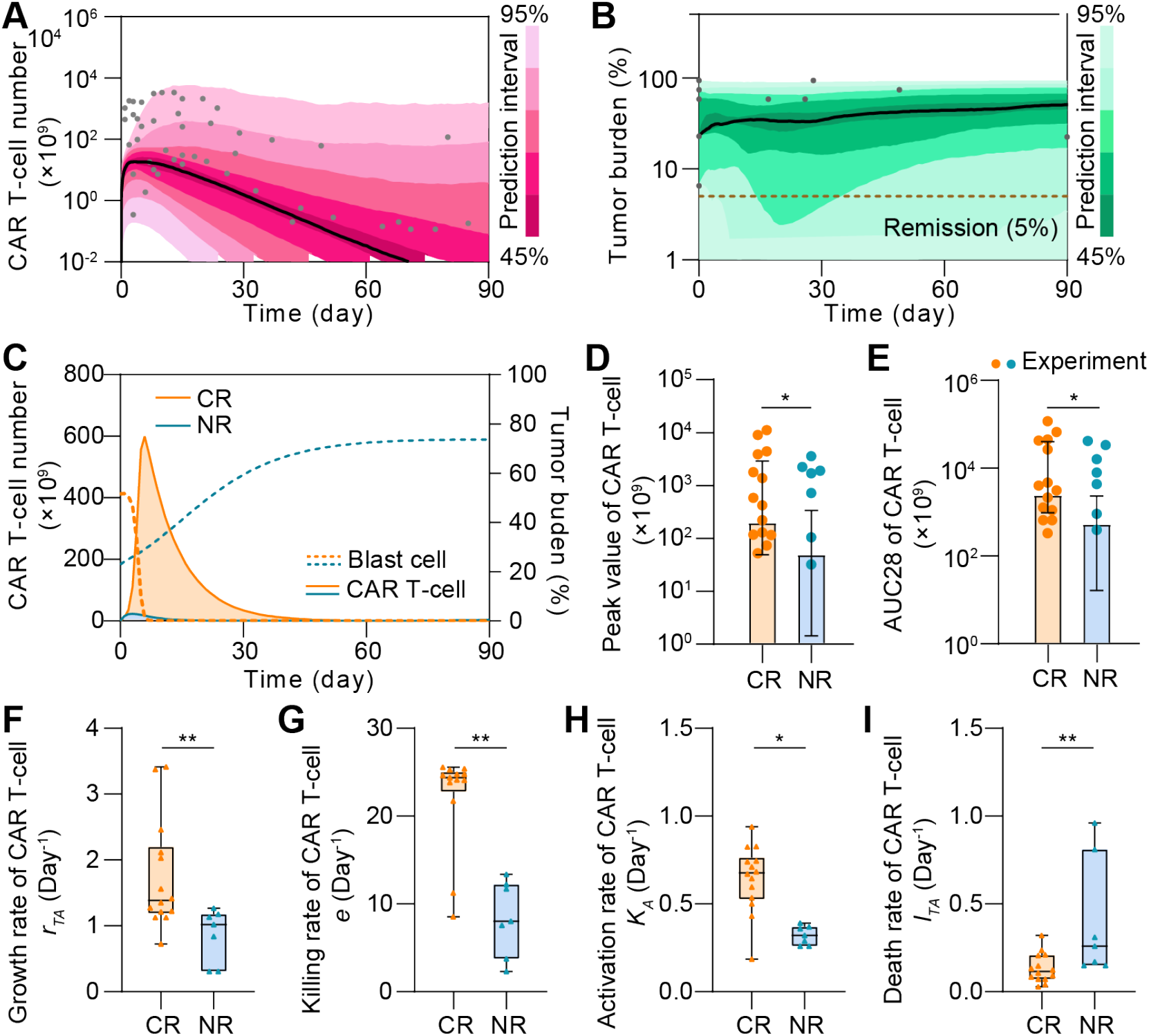
Comparative analysis of CR and NR patients *in silico*. (**A&B**) Calibration results of CAR T-cell (**A**) and tumor burdens (**B**) of NR patients with median (solid line) and 95% prediction interval (color bands). The dots represent the experimental data. (**C**) Variations of CAR T-cell and tumor burden of CR and NR patients. (**D&E**) Comparisons of the peak value (**D**) and AUC28 of CAR T-cell (**E**) between CR and NR patients. Bars are simulated results and dots are experiment results. Error bars are mean with 95% confidence interval. (**F-I**) The median growth rate of CAR T-cells of CR patients is 1.38 day^-1^, comparing with 1.02 day^-1^ of NR (**F**), the killing rate is 24.25 day^-1^ vs. 8.03 day^-1^ (**G**), the activation rate is 0.70 day^-1^ vs. 0.32 day^-1^ (**H**), and the apoptosis rate is 0.12 day^-1^ vs. 0.26 day^-1^ (**I**). Whiskers of boxplots are min to max value. *P*-values were calculated using Student’s or Welch’s t-test. *p <0.05, **p<0.01.

### *In silico* modeling reveals distinct CAR T-cell patterns in CD19+ and CD19^-^ relapse scenarios

In addition to those achieving continuous remission and showing non-response, some patients showed either CD19^+^ or CD19^-^ relapse after CAR treatment (6). To understand the heterogeneity between these two main relapse scenarios, we recalibrated the model with respective clinical data (**Figure 4A&B, Supplemental Figure S4&S5A-C, Supplemental Table S3&S4**). As shown by the individual level-calibration results, our computational model successfully mimicked the relapse progress at different days of relapse (**Figure 4C&D**). Furthermore, we found that the day of CD19^+^ relapse is positively related to the AUC28 of CAR T-cell (**Figure 4E**), whereas this is not the case for CD19^-^ relapse scenario (**Figure 4F**). To better explain this distinct pattern, we defined CAR T-cell function factor *F*_T_,

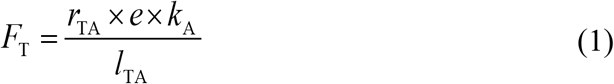

to quantitatively describe the overall CAR T-cell functionality consisting of growth, cytotoxicity and persistence where *r*_TA_ is the growth rate, *e* is the killing rate, *k*_A_ is the activated rate, and *l*_TA_ is the apoptosis rate of CAR T-cell. Simulation results demonstrated that the increase of *F*_T_ prolonged the day of CD19^+^ relapse (**Figure 4G**), but not that of CD19^-^ relapse (**Supplemental Figure S5D**). Such difference can be partially explained by the presence of CD19^-^ B-ALL cells before infusion of CAR T-cell and the following selective pressure by CAR T-cell (*4*). Besides, the genetic mutation causing loss of surface expression of CD19 and the bystander killing of CAR T-cell may further determined the progression of CD19^-^ B-ALL cells (*16, 17*). To integrate these two effects, we therefore defined the negative relapse factor *F*_NegR_,

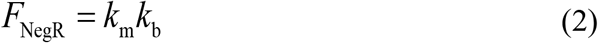

where *k*_m_ is the mutation factor considering the rate of gene mutation like alternative slicing with loss of exon 2 and the probability of lineage switch, and *k*_b_ is the bystander killing scaling factor depicting the killing efficacy of CAR T to CD19^-^ tumor cells comparing with to CD19^+^ ones. As a result, *F*_NegR_ demonstrated a high correlation with the day of CD19^-^ relapse, confirming its validity in depicting the mechanism of CD19^-^ relapse (**Figure 4H**). Together, our results reveal a distinct pattern of relapse across the CD19^+^ and CD19^-^ scenarios.

**Figure 4.**
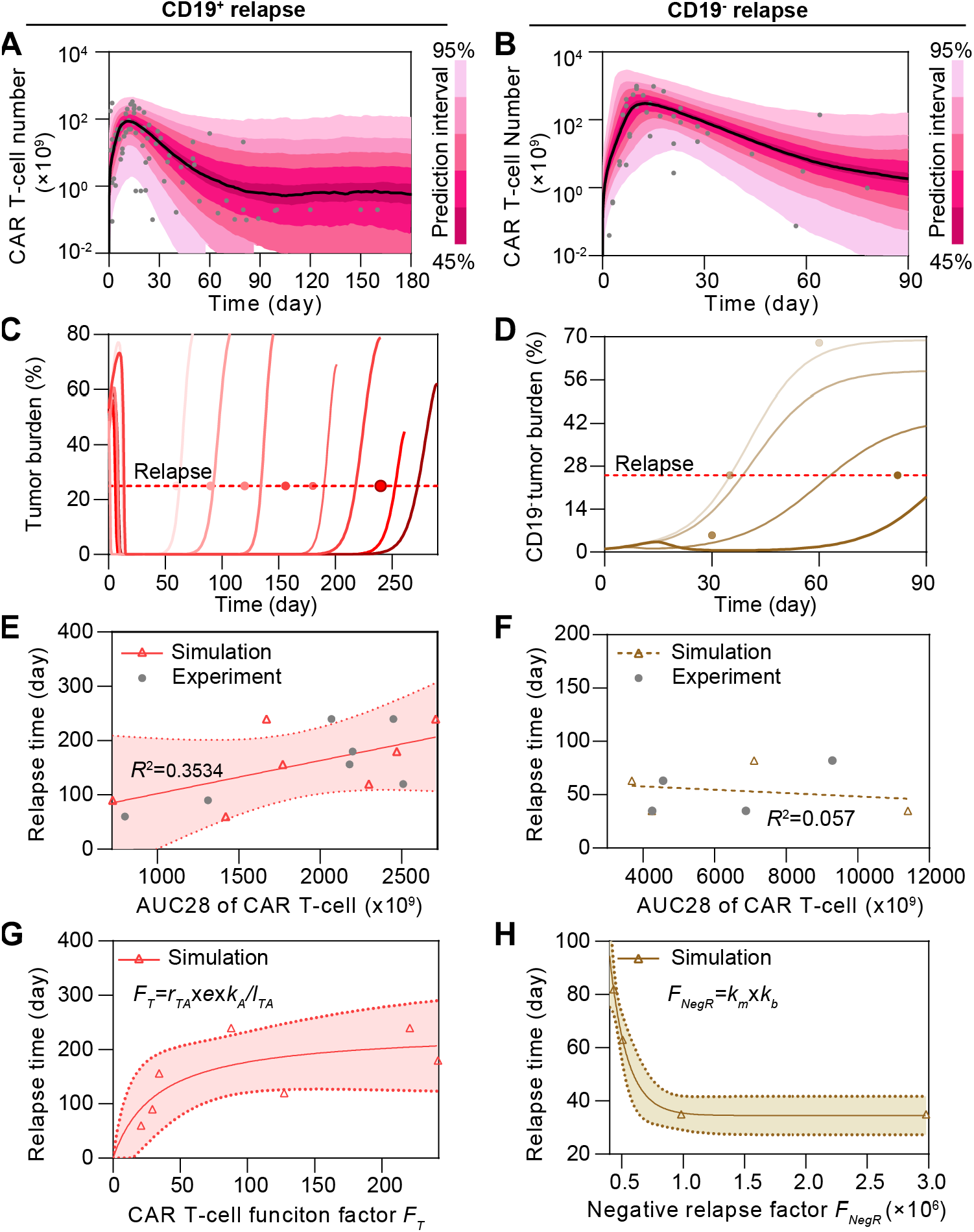
Comparative analysis of CD19^+^ and CD19^-^ relapse to CAR T-cell therapy *in silico*. (**A&B**) Calibration results of CAR T-cell of CD19^+^ (**A**) and CD19^-^ (**B**) relapse with median (solid line) and 95% prediction interval (color bands). (**C&D**) Individual fitting results of tumor burden of CD19^+^ (**C**) and CD19^-^ relapse (**D**). In A-D, the dots represent the experimental data and the lines represent the fitted curves. (**E&F**) Relationships between AUC28 of CAR T-cell and day of CD19^+^ (**E**) and CD19^-^ (**F**) relapse. (**G**) Variation of the day of CD19^+^ relapse as CAR T-cell function factor *F*_T_ changes. (**H**) Variation of the day of CD19^-^ relapse as negative relapse factor *F*_NegR_ changes. The band in E, G, H represents the 95% confidence interval.

### Key determinants underlie heterogeneous responses to CAR T-cell therapy

To understand the key factors determining the heterogeneous responses to CAR T-cell therapy, we systematically and comparatively analyzed the clinical data, such as the initial tumor burden and peak value of CAR T-cell in CR, NR, CD19^+^ relapse and CD19^-^ relapse patients after CAR T-cell therapy. There was no significant difference (P≥0.398) was observed in initial tumor burden among patients with different responses (**Supplemental Figure S6A**), suggesting that using initial tumor burden alone cannot determine CAR T-cell response. In parallel, we calculated the peak value and AUC28 of CAR T-cell for the four kinds of responses at the population level and found that CR and CD19^-^ relapse patients demonstrated higher values than did CD19^+^ relapse patients, whereas NR patients showed the minimum values (**Figure 5A**). We then tuned the values of the individual parameters (*r*_TA_, *e, k*_A_, and *l*_TA_) of CAR T-cell function factor *F*_T_ *in silico* and found that as expected the *F*_T_ parameter regulated the therapeutic effect of CAR T-cell treatment (**Supplemental Figure S6B&C**). These results together indicated the potential of *F*_T_ in differentiating these outcomes, such as CR, CD19^+^ relapse and NR after CAR T-cell therapy, though this was not the case for CD19^-^ relapse (**Figure 5B**). Furthermore, we scaled the population level calibrated parameters in *F*_T_ of different responses (±25% for CR, ±10% for CD19^+^ relapse, ±5% for NR), which again confirmed the usefulness of *F*_T_ (**Figure 5C**). Based on the clinical response and the distribution of *F*_T_ of patients (**Figure 5B**), the probability distribution of *F*_T_ of different responses were fitted with the MATLAB Distribution Fitter Toolbox (**Supplemental Figure S6D**). According to such probability distribution, the occurrence probabilities of CR, CD19^+^ relapse or NR at certain *F*_T_, i.e., *P*_*i*_ (*F*_T_) can be determined as

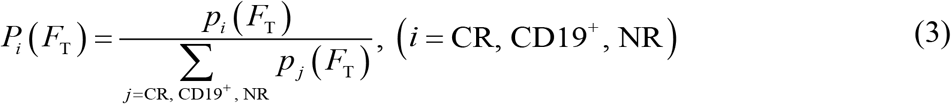

where *p* is the probability distribution of different responses (**Supplemental Figure S6C**). We found that as *F*_T_ increased, the most likely response to CAR T-cell therapy changed from NR to CD19^+^ relapse, and further to CR (**Figure 5D**). Besides, the distribution of scaled *F*_T_ (**Figure 5C**) fell into the range of corresponding response (**Figure 5D**). Similarly, since *F*_T_ cannot tell the characteristic of CD19^-^ relapse, we found instead that *F*_NegR_ determined the efficacy of CAR T-cell therapy in the scenario of CD19^-^ relapse (**Figure 5E**) and *in silico* experiments of changing *F*_NegR_ (by scaling *k*_m_ and *k*_b_) again confirmed such observation (**Figure 5F**). Together, our model help to identify critical determinants underlie heterogeneous responses to CAR T-cell therapy.

**Figure 5.**
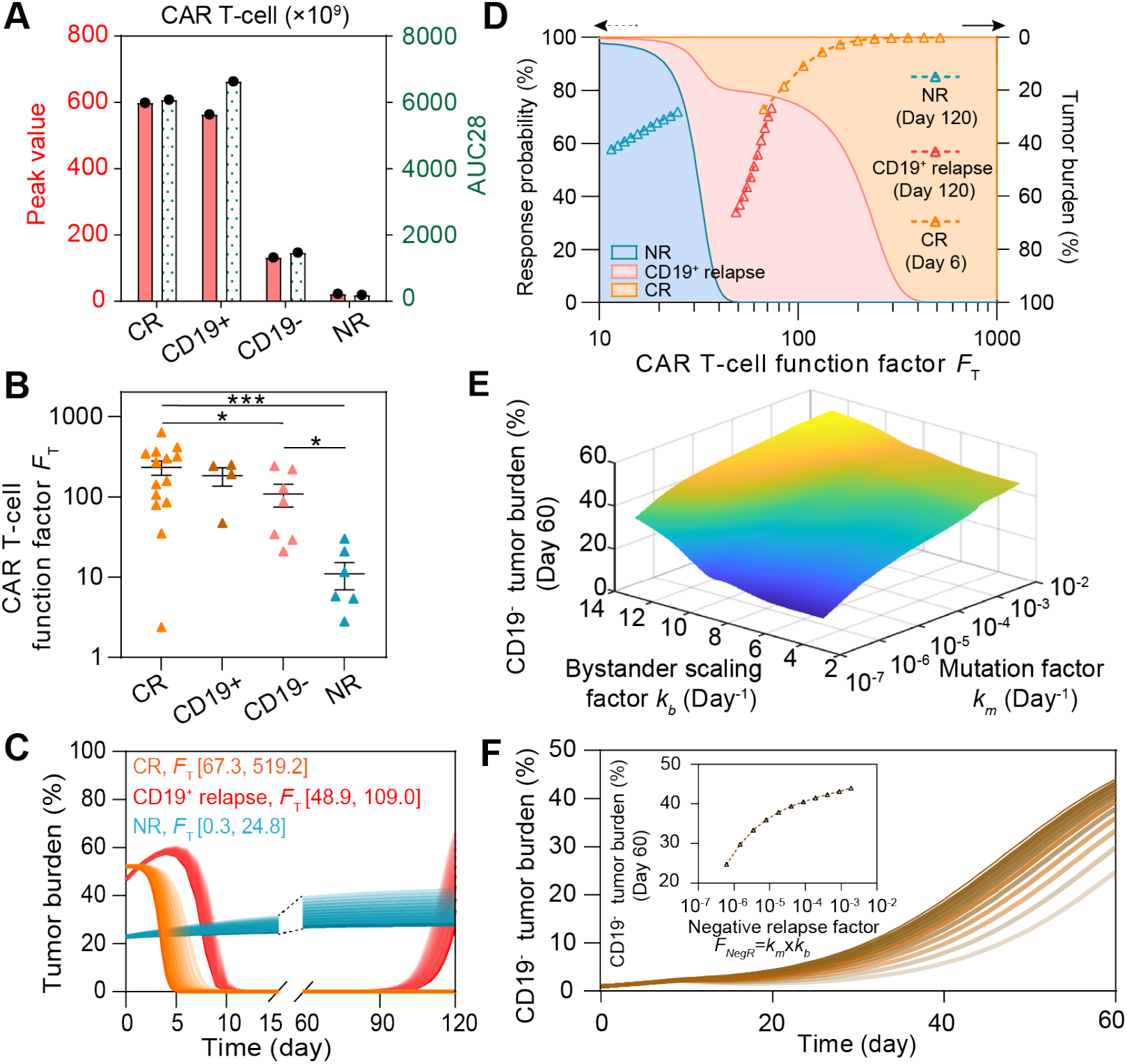
Key determinants in regulating CAR T-cell therapy response. (**A**) Comparisons of the peak value and AUC28 of CAR T-cell, and (**B**) CAR T-cell function factors *F*_T_ across patients of different responses. Error bars in B are mean ± stand error of mean (SEM). (**C**) Variations of tumor burden as *F*_T_ changes. (**D**) Variation of the response probability as *F*_T_ changes (left y-axis) and tumor burden under *F*_T_ in **C** (right y-axis). For a certain *F*_T_, the response probabilities of different responses are represented by different colors (blue for NR, red for CD19^+^ relapse, orange for CR) on the direction of y-axis, the sum of the response probabilities is 100%. As *F*_T_ changes, areas with different colors are generated. (**E**) Variations of CD19^-^ tumor burden as the bystander scaling factor *k*_b_ and mutation factor *k*_m_ change. (**F**) Variations of CD19^-^ tumor burden as the negative relapse factor *F*_NegR_ changes. *P*-values were calculated using Welch’s t-test. *p <0.05, ***p<0.001.

### *In silico* prediction of late response at early stage of CAR T-cell treatment

Prediction of patients’ late response to CAR T-cell therapy during early treatment stage will greatly improve patient outcomes by guiding the following regimen, especially for those with acute disease progression (*18-20*). Having demonstrated that our computational model accurately recapitulated the cellular dynamics of CAR T-cell therapy, we found that the response can be differentiated by the actual function level and dynamics of CAR T-cell in individual patients. Thus, we hypothesized that such responses can be predicted using our computational immuno-oncology model with input of clinically measurable and available patient information related to CAR T-cell dynamics, such as the peak value and AUC7 (area under the curve from day 0 to day 7) of CAR T-cell, at early stage of CAR T-cell treatment.

We first mapped the real-time results of four typical patient groups with different responses (**Figure 6A**). Unexpectedly, neither of peak value and AUC7 index alone demonstrated statistical differences among different groups, implicating that a single parameter is not proficient for clinical prediction (**Supplemental Figure S7**). Thus, we considered combining these two indices and defined the prediction factor *F*_P_ as 

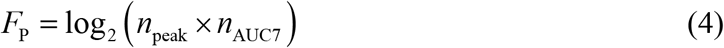

where *n*_peak_ is the peak value of CAR T-cell and *n*_AUC7_ is its AUC7 value. For different responses, we calculated *F*_P_ based on calibrated results of individual patients, and found that *F*_P_ showed statistical significance across CR, NR, and CD19^+^ relapse groups (**Figure 6B**). The results showed that early-stage CAR T-cell dynamics of CR and CD19^-^ relapse within the first month were similar in terms of the peak value, AUC7 of CAR T-cell and *F*_P_, as compared with NR and CD19^+^ relapse. Considering the similarity of CAR T-cell dynamics at early stage between CR and CD19^-^ relapse, these two response groups were combined together in the prediction and defined as CD&CD19^-^ relapse response.

**Figure 6.**
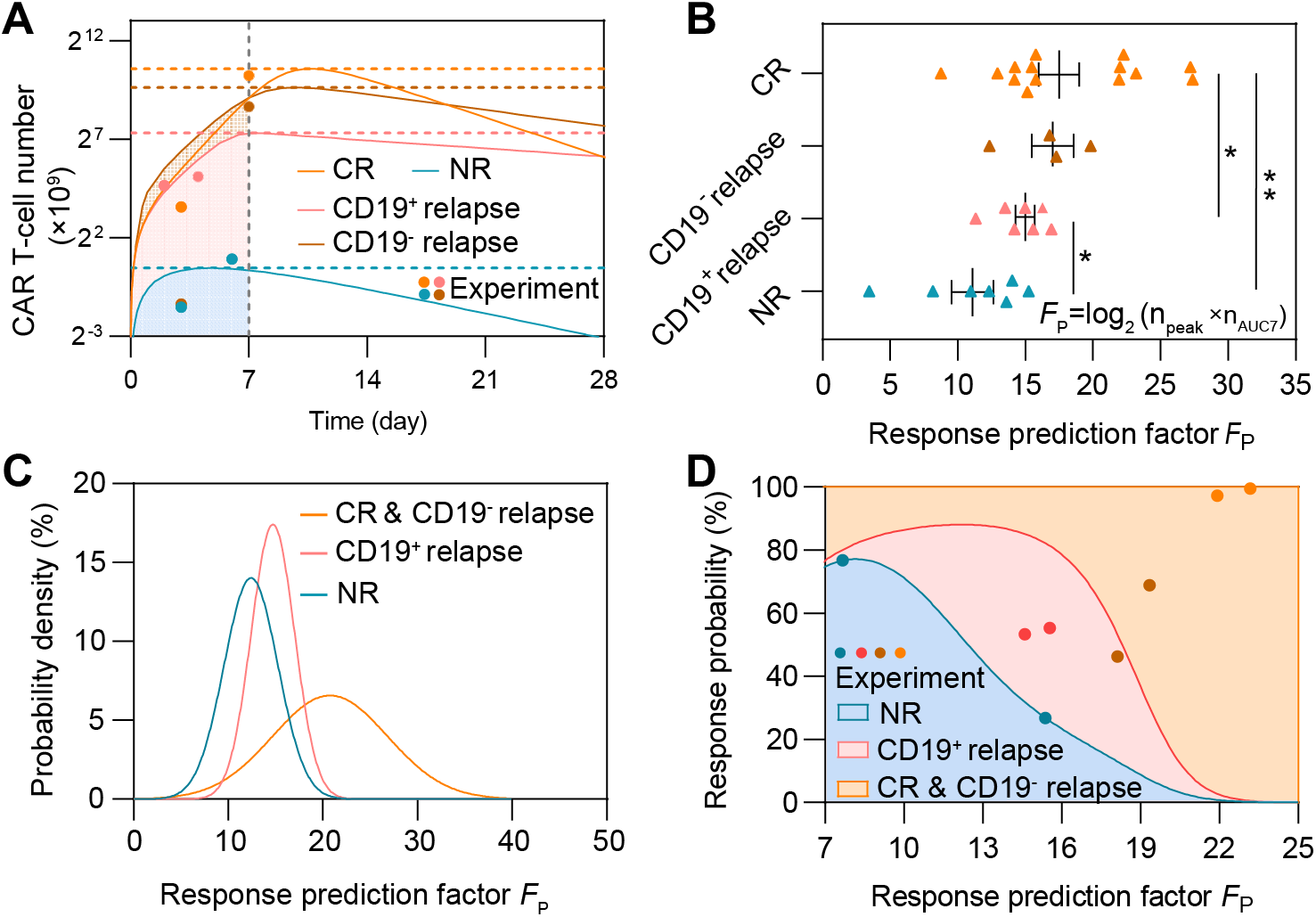
Prediction of CAR T-cell therapy response based on the peak value and AUC7 of CAR T-cells. (**A**) Variations of CAR T-cell number of typical patient groups with different responses (Group 8 of CR, Group 1 of CD19^-^ relapse, Group 7 of CD19^+^ relapse, and Group 1 of NR). (**B**) Comparisons of the response prediction factor *F*_P_. Error bars are mean ± SEM. (**C**) Probability density of different responses as *F*_P_ changes. (**D**) Variations of response probability as *F*_P_ changes. For a certain *F*_T_, the response probabilities of different responses are represented by different colors (blue for NR, red for CD19^+^ relapse, orange for CR&CD19^-^ relapse) on the direction of y-axis, the sum of the response probabilities is 100%. As *F*_T_ changes, areas with different colors are generated. The blue, red, brown, and orange dots represent experimental results of NR, CD19^+^ relapse, CD19^-^ relapse and CR patients respectively to validate prediction ability. *P*-values were calculated using Student’s or Welch’s t-test. *p <0.05, **p<0.01.

To further validate the prediction ability of *F*_P_, for CR&CD19^-^ relapse, we chose 7 patients for calibrating the relationship between *F*_P_ and the response, and 4 (2 CR, 2 CD19^-^ relapse) for prediction; for NR and CD19^+^ relapse, we chose 5 for calibration and 2 for prediction. The probability distributions of *F*_P_ of different responses are fitted with the MATLAB Distribution Fitter Toolbox (**Figure 6C**). Based on the probability distribution, the occurrence probabilities of each response at certain *F*_P_, i.e., *P*_*i*_ (*F*_P_) can be determined as

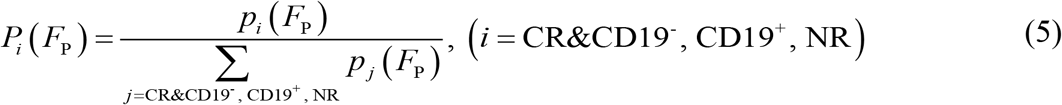

where *p* is the probability distribution of different responses. We found that as *F*_P_ increased, the most likely response of CAR T-cell therapy changed from NR to CD19^+^ relapse, and then to CR&CD19^-^ relapse (**Figure 6D**). *F*_P_ of each patient for prediction was calculated and the corresponding probabilities of different responses can be determined (**Figure 6D**). As a result, *F*_P_ of two NR patients are 15.4 and 7.7 and the predicted probabilities of non-response are of 26.7% and 76.7%; *F*_P_ of two CD19^+^ relapse patients are 14.6 and 15.5 and the predicted probabilities of CD19^+^ relapse are 53.3% and 55.3%; *F*_P_ of two CD19^-^ relapse patients are 18.1 and 19.3 and the predicted probabilities of CR&CD19^-^ relapse are 46.3% and 68.9%; *F*_P_ of two CR patients are 21.9 and 23.2 and the predicted probabilities of CR&CD19^-^ relapse are 97.2% and 99.5%.

Given that *F*_P_ prediction depended on the peak value of CAR T-cell which requires clinical monitoring of patients up to 2 weeks, we sought to predict patient outcome with only input of the first 7-day clinical data of CAR T-cell dynamics and the initial tumor burden of a given patient. We then calibrated the computational model based on the early-stage CAR T-cell dynamics (**Equations 6-8**) and obtained the subsequent time-series results of CAR T-cell and tumor cells (**Figure 7A-C**). To validate the feasibility of the proposed method, we first tested one representative patient for different response group and found that the responses of clinical patient cohort can be correctly predicted (**Figure 7A-C**).

**Figure 7.**
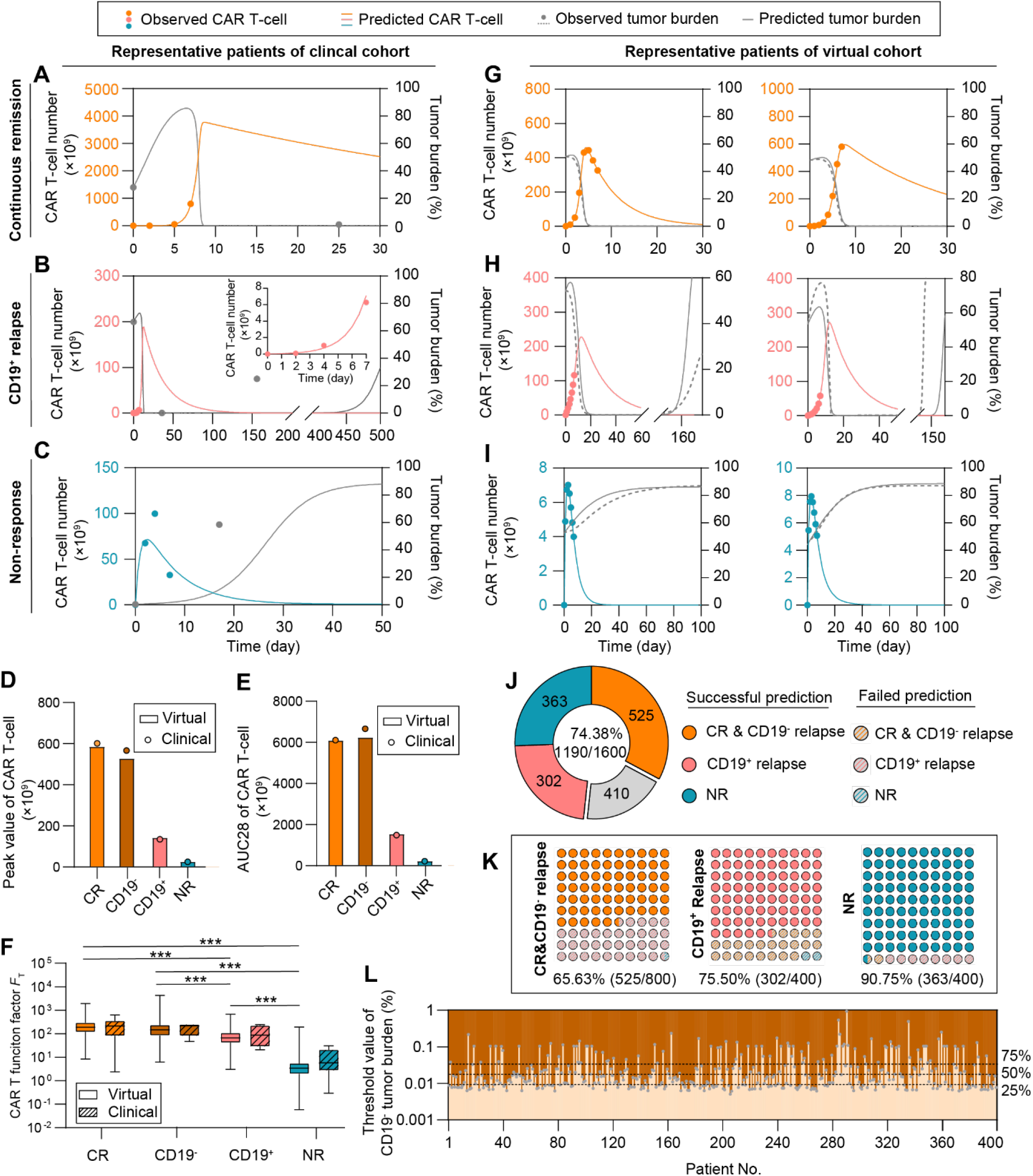
Prediction of patient responses to CAR T-cell therapy based on the CAR T-cell dynamics within first 7 days of treatment. (**A-C**) Prediction results based on clinical patients of CR (**A**), CD19^+^ relapse (**B**), and NR (**C**). The minor graph in B shows CAR T-cell dynamics observed and predicted in the first 7 days in a smaller y-axis scale for better display. (**D**) Median peak value and (**E**) AUC28 of CAR T-cell of virtual cohort patients and comparisons with clinical cohort patient data. (**F**) CAR T-cell function factors of virtual patients and comparisons with clinical data. No significant differences between virtual and clinical patients of the same response, and between CR and CD19^-^ relapse patients. Whiskers of boxplots are min to max value. (**G-I**) Real-time prediction results based on virtual patient cohorts. The prediction method predicted the CR (**G**), CD19^+^ relapse (**H**) and NR (**I**) of CAR T-cell therapy as observed. (**J**) Overall prediction accuracy (74.38%). (**K**) Prediction accuracy of different response: 65.63% for CR & CD19^-^ relapse, 75.50% for CD19^+^ relapse, and 90.75% for NR. (**L**) Threshold values of initial CD19^-^ tumor burden to induce CD19^-^ relapse. Dotted lines indicate the values of initial CD19^-^ tumor burden with the occurrence probability (25%, 50%, 75%) of CD19^-^ relapse obtained from population level statistics. *P*-values were calculated using Student’s or Welch’s t-test. ***p <0.001.

To test the accuracy of our prediction method on a larger scale with higher reliability, we next generated clinical-derived virtual patient cohorts for a clinical trial simulation to complement the present clinical data which is of small quantity and density (Details see ‘Generating virtual patient cohorts’ in **Methods**). In general, 400 virtual patient data points per CR, CD19^-^ relapse, CD19^+^ relapse and NR cohorts were generated (**Supplemental Table S5-8**), based on Gaussian distribution of the population level parameters calibrated from present clinical patient cohort data (**Table 1**). To validate the applicability of those virtual patient cohorts, we first compared the peak value (**Figure 7D**) and AUC28 (**Figure 7E**) of CAR T-cells between virtual and clinical patient cohorts of different responses (**Figure 5A**). The results showed good consistency in both absolute and relative values, indicating the quantity and quality of virtual patient cohorts matched the clinical ones. We further calculated *F*_T_ of virtual patient cohorts and found no significant differences between clinical patient cohorts and virtual patient cohorts (**Figure 7F**), again confirming the usability of virtual patient cohorts. As a result, our clinical trial simulation with virtual patient cohorts successfully reproduced real-time results of corresponding clinical patient cohorts and predicted patient responses (**Figure 7G-I, Supplemental Figure S8**-**11**). Overall, our prediction method reached a prediction accuracy of 74.38% in total of 1,600 virtual patients with respective prediction accuracy of 65.63% (525/800) for CR&CD19^-^ relapse, 75.50% (302/400) for CD19^+^ relapse, and 90.75% (363/400) for NR (**Figure 7J&K**). For CR and CD19^-^ relapse, CAR T-cell dynamics at early stage are similar hence their responses were combined as CR&CD19^-^ relapse (**Figure 7A, Supplemental Figure S12**). The high prediction accuracy of NR can be partially explained by the lower differences between the predicted and observed values of *F*_T_, Δ*F*_T_ (**Supplemental Figure S13**).

**Table 1.** Estimated parameters and initial values of CAR T-cell and B-ALL cell.

| Parameter | Description | Unit | CR* | NR* | CD19<sup>+</sup><br>relapse | CD19<sup>-</sup><br>relapse | Reference |
| --- | --- | --- | --- | --- | --- | --- | --- |
| $r_P$ | Growth rate of CD19<sup>+</sup> B-ALL cell | Day<sup>-1</sup> | 0.069 | 0.080 | 0.21 | 0.071 | (45, 46) |
| $r_{TA}$ | Growth rate of activated CAR T-cell | Day<sup>-1</sup> | 1.62 | 0.99 | 0.99 | 1.5 | (13, 47) |
| $l_{TA}$ | Apoptosis rate of activated CAR T-cell | Day<sup>-1</sup> | 0.12 | 0.55 | 0.12 | 0.11 | (48, 49) |
| $l_{TN}$ | Apoptosis rate of non-activated CAR T-cell | Day<sup>-1</sup> | 3e-5 | 9.2e-4 | 6e-7 | 2e-7 | |
| $n_C$ | B-ALL cell carrying capacity | | 2939.1 | 6101.58 | 19877.4 | 2585.74 | |
| $e$ | Killing rate of activated CAR T-cell | Day<sup>-1</sup> | 22.72 | 6.58 | 20.31 | 19.34 | (10) |
| $K_P$ | Saturation constant to CAR -T cell killing rate | | 5891.5 | 7067.07 | 1050.19 | 11040.05 | |
| $K_r$ | Saturation constant to CAR T-cell growth rate | | 637.64 | 3431.65 | 1983.64 | 1360.54 | |
| $K_A$ | Saturation constant to CAR T-cell activation rate | | 1808.0 | 0.0052 | 54.68 | 11883.73 | |
| $k_A$ | Activation rate of CAR T-cell | Day<sup>-1</sup> | 0.65 | 0.31 | 0.44 | 0.58 | |
| $r_N$ | Growth rate of CD19<sup>-</sup> B-ALL cell | Day<sup>-1</sup> | / | / | / | 0.1 | (45, 46) |
| $k_m$ | Mutation factor | Day<sup>-1</sup> | / | / | / | 1.5e-7 | (50) |
| $k_b$ | Bystander killing scaling factor | | / | / | / | 7.9 | |
| $K_N$ | Saturation rate of CD19<sup>-</sup> B-ALL cell to killing efficacy | | / | / | / | 16956.03 | |
| $n_{P0}$ | Initial value of CD19<sup>+</sup> B-ALL cell | ×10<sup>9</sup> cells | 2200.24 | 589.676 | 1764.25 | 1467.01 | |
| $n_{N0}$ | Initial value of CD19<sup>-</sup> B-ALL cell | ×10<sup>9</sup> cells | / | / | / | 19.89 | Assumed (1%) |
| $n_{TN0}$ | Initial value of non-activated CAR T-cell | ×10<sup>9</sup> cells | 16.5 | 71.45 | 12.26 | 8.97 | |

\*CR represents continuous remission and NR represents non-response.

To further differentiating CR and CD19^-^ relapse patients in the group of CR&CD19^-^ relapse response, we chose the initial CD19^-^ tumor burden as an index to predict the occurrence of CD19^-^ relapse (**Equation 9**). Parameters designated to the model are based on the calibration of CR&CD19^-^ relapse response combining with parameters related to CD19^-^ relapse (**Table 1**). The response varies from CR to CD19^-^ relapse by increasing the initial CD19^-^ tumor burden, so its threshold value of each individual and population level results with certain occurrence probabilities to induce CD19^-^ relapse can be determined (**Figure 7L, Supplemental Figure S14**). These results together confirm the prediction power of our computational CAR T-cell immuno-oncology model, which can provide guidance for clinical treatment and regimen.

## Discussion

Computational models of immunotherapy provide a valuable tool for *in silico* clinical modeling and patient stratification (*6, 7*). In the present study, we constructed a mathematical model of the critical leukemia-immune interactions with clinical dataset, determined key factors effecting treatment efficacy, and predicted patient response to CAR T-cell therapy with early-stage CAR T-cell dynamics data. We systematically explored that the dynamic interactions between B-ALL and CAR T-cell and found that CAR T-cell function index *F*_T_ inferred patients’ outcomes ranging from CR, NR to CD19^+^ relapse. By contrast, a negative relapse index *F*_NegR_ including the mutation rate and the bystander killing rate to CD19^-^ B-ALL cells determined the probability and day of relapse for CD19^-^ relapse cases. Applying this computational model, we were able to define the prediction factor *F*_P_ with the early-stage CAR T-cell dynamics data including the peak and AUC7 values of CAR T-cell to predict CAR T-cell treatment outcomes for individual patients.

Comparing with many other models only end-point data are provided, our model predicted the prognosis of patients with time series data. Many present methods for response prediction required detection of certain prognostic biomarkers, for example inducible COStimulator (ICOS), CD27^+^PD-1^−^CD8^+^ T cell population, lactate dehydrogenase (LDH), and C-reactive protein (CRP) (*21-24*), increasing the complexity in practical operation. More importantly, current biomarkers failed to accurately and timely predict the prognosis of patients who underwent CAR T-cell therapy. By contrast, our prediction method needs only inputs of the number of CAR T-cell at early stage of treatment, which is easy to access and commonly measured in clinic, making it acquire higher reliability and potential to be translated into real-world applications.

Computational models usually adopt clinical data from different literatures acquired by different measuring methods (e.g., flow cytometry, qPCR, and morphological testing) of different biopsies took from different tissue samples (e.g., saliva, peripheral blood, and bone marrow). Such a dataset possesses a large variance, making it difficult for model fitting and for obtaining one set of parameters that would work for all patients under different treatment conditions. Thus, unifying the data from different sources and formats before fitting *in silico* models is particularly important. Accordingly, we built up a unifying code to interpret the relationship between clinical data with different units coming from different body parts with assumptions (*12*) and data (*25*). This unifying process of clinical data highlights a novel way to accurately recapitulate the patient/clinical process, which previous strategies are limited to offer. Moreover, the accuracy and reliability of the prognostic model highly depend on the quantity and quality of clinical data to establish and calibrate it. Most critically, limited availability of clinical data prevents large-scale validation of computational models including ours. We therefore conducted clinical trial simulation studies expanding the patient cohorts from 209 to 1,600 individuals. The generation of virtual patient cohorts was based on calibration results of clinical patient cohorts, and the similarity of these two cohorts was carefully confirmed, both ensuring the credibility of clinical trial simulations. The proposed methods to unify and expand clinical data have been proven to be practical and feasible, but it has to be admitted that original clinical data with large amount and uniformed standard are optimal, necessitating collaboration with clinicians on designing the scheme of data collection before the start of clinical trials for a better validation and translation of the model. Moreover, the current model tried to fully utilize clinical data obtained from different trials with a uniform and reasonable standard, but it can be improved with more considerations of interpatient variability. For example, lymphodepletion pretreatment seems to effect CAR T-cell expansion (*26*) and second-generation CAR T-cell products with CD28 or 4-1BB costimulatory domains exhibit differences in magnitude and persistence (*27*) although both showed well treatment efficacy to B-ALL. Future extension of the computational model including these factors will develop its potential in exploring new insights.

Relapse scenarios, which were less discussed in current computational models, were included in our model. Our simulation results demonstrated a prediction of the prognosis of CD19^+^ relapse based on early-stage CAR T-cell dynamics data, but not that of CD19^-^ relapse. The clinical responses of CD19^-^ relapse patients were found very similar to CR patients at the early stage of CAR T-cell treatment. In our model, the index of initial CD19^-^ tumor burden demonstrated a good potential in predicting CD19^-^ relapse. However, such index of CD19^-^ tumor burden is current less focused and measured clinically, let alone the early-stage measurement (like until day 7) of them, which could provide more sufficient information to calibrate our model for real-world clinical prediction. Although it is clear that presence of CD19^-^ tumor cell population caused the CD19^-^ relapse (*28*) and our model identified the potential threshold of the CD19^-^ cell population for CD19^-^ relapse, the mechanisms causing the loss of surface expression of CD19 remain poorly understood. Thus, more insightful studies about the mechanism of CD19^-^ relapse will also enable us to include more factors in the model with improved knowledge to achieve a better description and prediction of CD19^-^ relapse.

In addition to the direct interactions between CAR T-cell and B-ALL cell, microenvironmental cues, for example, immune cells like myeloid-derived suppressor cells, regulatory T cells and tumor-associated macrophages, and their secreted immunosuppressive cytokines in the bone marrow, might explain the distinct functionalities of CAR T-cell for patients with different response (*29*). As demonstrated by our recent studies (*30*), an *in silico* modeling of the heterogeneous tumor microenvironments would be particularly valuable for dissecting and screening of immunotherapies. Incorporation of these immune cell components into our computational immuno-oncology model may help dissecting out these immunological mechanics involved in the tumor microenvironment affecting treatment outcomes, though it remains currently impractical due to highly limited clinical data. Computational models were also applied to study the cytokine release syndrome (CRS), an adverse side effect elicited by CAR T-cell therapy (*31, 32*). However, most of existing CRS models lack clinical data and failed to dissect actual cellular factors affecting the severity of CRS and corresponding treatments. We believe that modeling of cytokines with our computational CAR T-cell therapy model would help to determine the biological mechanisms and the risk to develop severer CRS and their potential relationships with the varied therapeutic outcomes.

In conclusion, we have established a computational immuno-oncology model of CAR T-cell therapy recapitulating key cellular dynamics observed in clinical trials, revealed key determinants of treatment efficacy, and predicted patient outcomes. We hope that this computational platform inputted with early-stage clinical data can improve the clinical outcomes and guide clinical trials of personalized CAR T-cell therapy.

## Materials and Methods

### Model construction

#### CD19^+^ B-ALL Cell

It is assumed that the proliferation of CD19^+^ B-ALL cell subjects to logistic growth (*33*), and they are eliminated by activated CAR T-cell as

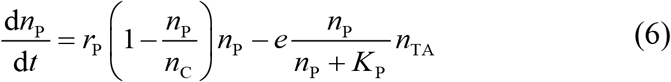

where *n*_P_ (P represents positive) is the number of CD19^+^ B-ALL cell, *n*_C_ is the carrying capacity of B-ALL cell in the tumor microenvironment, *n*_TA_ is the number of activated CAR T-cell, and *e* is their killing rate. The efficacy of the elimination of B-ALL cell follows Michaelis-Menten kinetics with a Michaelis constant *K*_P_, denoting the saturation effect from B-ALL cell to killing efficacy.

#### CAR T-cell Activation

CAR T-cell can kill B-ALL cell only after being activated by CD19^+^ B-ALL cell from initial non-activated status. The generation of activated CAR T-cell can be expressed as

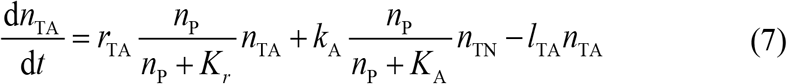

where *r*_TA_ is the growth rate of activated CAR T-cell, *k*_A_ is the activation rate from initial non-activated CAR T-cell, *n*_TN_ is the number of non-activated CAR T-cell, and *l*_TA_ is the apoptosis rate of activated CAR T-cell. The growth rate and activation rate of CAR T-cell are affected by CD19^+^ B-ALL cell with saturation constant *K*_*r*_ and *K*_A_ respectively.

Thus, the variation of non-activated CAR T-cell can be expressed as

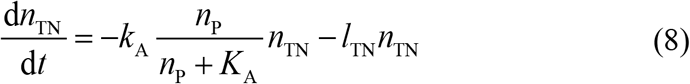

where the first term represents the conversion to activated status, and *l*_TN_ is the apoptosis rate of non-activated CAR T-cell.

#### CD19^+^ and CD19^-^ Relapse

For CD19^+^ relapse, CD19 antigen is still present on the surface of B-ALL cells and can be detected by flow cytometry, so the model of CD19^+^ relapse can still be described by equations (6)-(8). However, the response of CD19^+^ relapse is different from CR, and the key mechanism lies in the poor function (expansion, cytotoxicity, and persistence) of CAR T-cell (*34*). For CD19^+^ relapse patients, CAR T-cell parameters in the model such as growth rate, killing rate, activation rate and apoptosis rate should be inferior to CR patients (*35*).

In CD19^-^ relapse, there exist CD19 antigen absent B-ALL cells, causing tumor to evade CAR-mediated recognition and clearance in spite of CAR T-cell persistence (*4*). The mechanism of CD19 loss is attributed to immune pressure selection (CD19-negative tumor cells have already existed before CAR T-cell therapy) and CD19 gene mutation (e.g., alternative slicing with loss of exon 2) (*16*). In CD19^-^ relapse, although CAR T-cell cannot eliminate CD19^-^ tumor cell through antigen recognition, activated CAR T-cell can mediate tumor lysis against the antigen negative fraction in an antigen independent, cell-cell contact-mediated manner in the vicinity of the cells they’re designed to target, which is called bystander killing effect (*17*). So the variation of CD19^-^ tumor cells can be expressed as

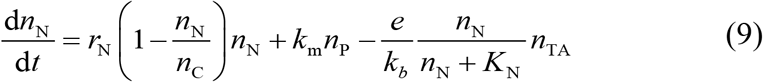

where *n*_N_ (N represents negative) is the number of CD19^-^ tumor cell, *k*_m_ is the mutation factor causing CD19 loss from CD19^+^ B-ALL cell, *k*_b_ is the bystander killing scaling factor to CD19^+^ killing efficacy, and *K*_N_ is the saturation constant depicting the effect of CD19^-^ B-ALL cells to bystander killing.

### Collecting and processing clinical data

#### Merging the individual data into groups

Real-time data of CAR T-cell and tumor burden including the information of 209 patients were collected from clinical studies (*25, 35-43*). For clinical trials that did not disclose raw data, the clinical data were digitized by WebPlotDigitizer (https://automeris.io/WebPlotDigitizer) from published figures of these clinical studies.

Although in some studies there are clinical data of individual patients, for some studies, individual data had been preprocessed and we can only get the statistical values like medians from such references. However, it does not weaken the reliability of the calibration process and the rationality of the computational model, since such statistical values still contain the information of individual patient. Thus, for one piece of available clinical data (containing one or several individuals), we called it a group. After summarizing the data in references, we got 32 groups (14 for CR, 7 for NR, 7 for CD19^+^ relapse, and 4 for CD19^-^ relapse) including the clinical information of 209 individuals (148 for CR groups, 24 for NR groups, 20 for CD19^+^ relapse groups, and 17 for CD19^-^ relapse groups) (**Supplemental Table 1-4**).

#### Converting data from peripheral blood to bone marrow

The computational model dissects the cellular behaviors in the B-ALL bone marrow during CAR T-cell therapy. However, samples are often collected in peripheral blood clinically (e.g., CAR T-cell). So, such data need to be converted into their equivalent values in bone marrow. For data acquired in peripheral blood, it is assumed that the average volume of blood sample is 5 mL and there are 5000 mL bloods in human body (*44*). So the number of cells in the whole peripheral blood can be calculated. After that, it is assumed that comparing with bone marrow, about 1% cells are in peripheral blood (*12*). Thus, the number of cells in bone marrow can be obtained.

#### Unifying the units of B-ALL cells

In most clinical studies, the unit of B-ALL cells is tumor burden (%). However, the proposed CAR T-cell therapy model is cellular-based, so for computational processes like calibration and simulation, the unit of B-ALL cells need to be converted into number of B-ALL cells. We got the clinical pre-treatment tumor burdens (**Table 1** of ref. (*25*)) and absolute numbers of B-ALL cells (**Figure 1G** of ref.(*25*)) of MRD^-^ patients, and their relationship was determined after fitting (**Supplement Figure S15**). The expression of the fitted results is

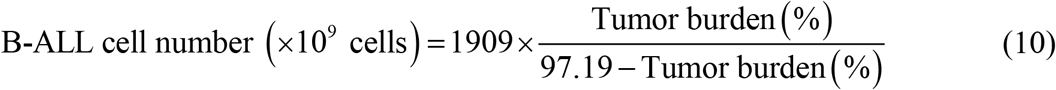

It is noted that although the unit of B-ALL cells were converted into absolute numbers for computation, the unit was converted back into tumor burden (%) for presentation in our paper to make it more understandable and compliant with clinical custom.

#### Unifying the units and scaling of CAR T-cells

Like B-ALL cells, the units of CAR T-cell were all converted into number of CAR T-cells (×10^9^) for computation. Clinically the amount of CAR T-cell are often measured by quantitative PCR (qPCR) (normally in the unit of copies CAR/μg DNA) in peripheral blood. From **Figure 1F** of ref. (*25*), we obtained the qPCR values and corresponding absolute numbers of CAR T-cell of MRD^-^ patients. Considering the samples of these two measurement methods were of different numbers (n=14 for qPCR values and n=11 for absolute numbers) in (*25*), we got the median of these two values and determined their relationship as

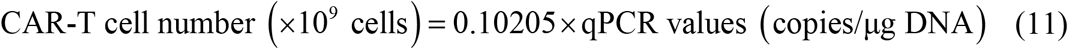

Comparing to CR patients and NR patients, clinical data of CAR T-cell of CD19^+^ and CD19^-^ relapse is relatively lack in clinical studies, thus requiring more process.

According to current clinical researches, CD19^+^ relapse is associated with loss of CAR T-cell function. In contrast, CD19^-^ relapse occurs despite CAR T-cell functional persistence and they are likely to be independent of CAR T-cell parameters per se (*34*). For CD19^+^ relapse patients, the function of CAR T-cell is between CR and NR patients; and for CD19^-^ relapse patients, the function of CAR T-cell is similar to CR patients (*35*). So it is reasonably assumed that the values of CAR T-cell of CD19^+^ relapse patients is between CR and NR patients, of CD19^-^ relapse patients is close to CR patients. In this paper, based on the above assumption, we scaled the limited amount of data of CAR T-cell for CD19^+^ and CD19^-^ relapse. With the support of biological and clinical studies, the reliability of the computational model is maintained although with scaling, especially for comparisons and analysis inside CD19^+^ or CD19^-^ relapse patient groups.

### Parameter estimation

Parameter estimation (on individual and population level) were conducted using the stochastic approximation expectation maximization (SAEM) algorithm for nonlinear mixed-effect modeling (NLME), as implemented in Monolix version 2020R1 (Lixoft, France). The estimated population level-parameters and initial values related to CAR T-cells and tumor cells (**Table 1**) were obtained. References in **Table 1** determined the rational ranges of estimated parameters.

### Generating virtual patient cohorts

The parameters of virtual patient cohorts were generated obeying the Gaussian distribution with mean value *μ* equaling to the population leveled-parameters (**Table 1**) of CR, CD19^+^ relapse and NR patients. The standard deviation equals to 1/3 *μ* ensuring the parameters are larger than 0. For each population leveled-parameters of each response, we generated 400 sets of parameters of continuous remission, CD19^-^ relapse, CD19^+^ relapse and non-response respectively.

### Statistics

Data were first analyzed for normality and then compared with unpaired Student’s t test or Welch’s t test using Prism 7.0 (GraphPad). p<0.05, p<0.01, and p<0.001 were considered significantly different. The results, including the error bars in the graphs, were given as the mean ± standard error of mean (SEM) or boxplot with whisker of min to max value. Details are reported in each figure.

## Data Availability

All the relevant data are included in the manuscript and its supplementary material file. The code for the computational model is available on Github (https://github.com/Lunan12/CAR-T-model).

## Author contributions

C.M. and W.C. conceived the project; L.L. and Z.Z. constructed the computational model and performed experiments; L.L., Z.Z., and C.M. analyzed data; L.L., Z. Z., C.M. and W.C. wrote the manuscript; W.C. supervised the project. All authors edited and approved the final manuscript.

## Supplementary Information

**Table S1.** Merged clinical data of continuous remission patient groups from individuals.

| Group | Reference | Number of individuals | Statistical value |
| --- | --- | --- | --- |
| 1 | (35) | 45 | LOESS (local polynomial regression) curve fitting |
| 2 | (25) | 14 | First quartile |
| 3 | (25) | 14 | Second quartile |
| 4 | (25) | 14 | Third quartile |
| 5 | (36) | 59 | Median & geometric mean |
| 6 | (37) | 1 | Original (Patient 1) |
| 7 | (38) | 8 | Mean |
| 8 | (39) | 1 | Original (No. MSK-ALL05) |
| 9 | (40) | 4 | BM M1 status (<5%) |
| 10 | (40) | 4 | BM M2 status (5-25%) |
| 11 | (40) | 9 | BM M3 status (>25%) |
| 12 | (41) | 1 | Original (No. MSK-ALL13) |
| 13 | (41) | 1 | Original (No. MSK-ALL14) |
| 14 | (41) | 1 | Original (No. MSK-ALL13) |
| Total |  | 148 |  |

**Table S2.**
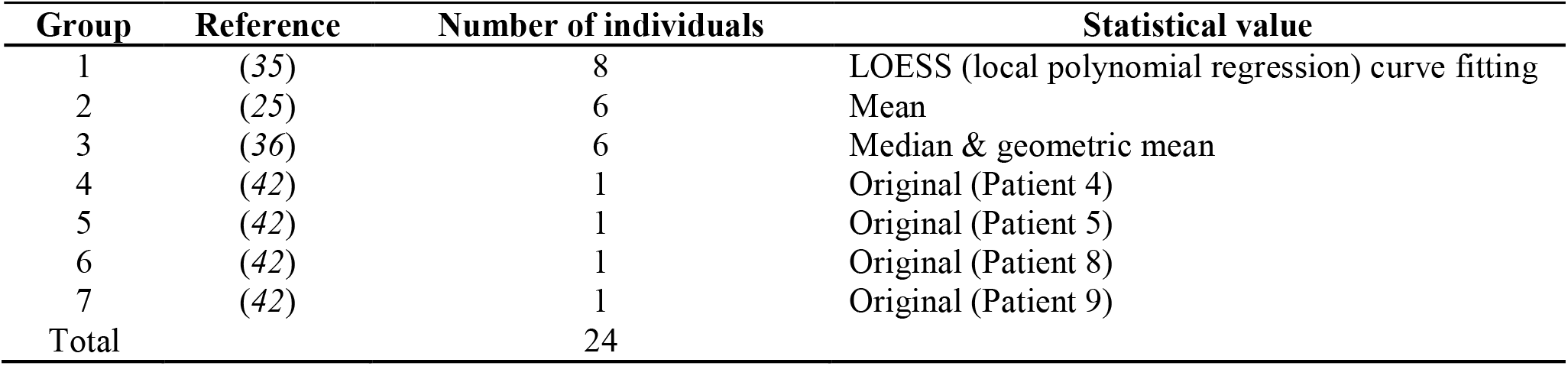
Merged clinical data of non-responding patient groups from individuals.

| Group | Reference | Number of individuals | Statistical value |
| --- | --- | --- | --- |
| 1 | (35) | 8 | LOESS (local polynomial regression) curve fitting |
| 2 | (25) | 6 | Mean |
| 3 | (36) | 6 | Median & geometric mean |
| 4 | (42) | 1 | Original (Patient 4) |
| 5 | (42) | 1 | Original (Patient 5) |
| 6 | (42) | 1 | Original (Patient 8) |
| 7 | (42) | 1 | Original (Patient 9) |
| Total |  | 24 |  |

**Table S3.** Merged clinical data of CD19^+^ relapse patient groups from individuals.

| Group | Reference | Number of individuals | Statistical value |
| --- | --- | --- | --- |
| 1 | (35) | 14 | LOESS (local polynomial regression) curve fitting |
| 2 | (39) | 1 | Original (No. MSK-ALL04) |
| 3 | (42) | 1 | Original (Patient 1) |
| 4 | (42) | 1 | Original (Patient 2) |
| 5 | (42) | 1 | Original (Patient 3) |
| 6 | (42) | 1 | Original (Patient 6) |
| 7 | (42) | 1 | Original (Patient 7) |
| Total |  | 20 |  |

**Table S4.**
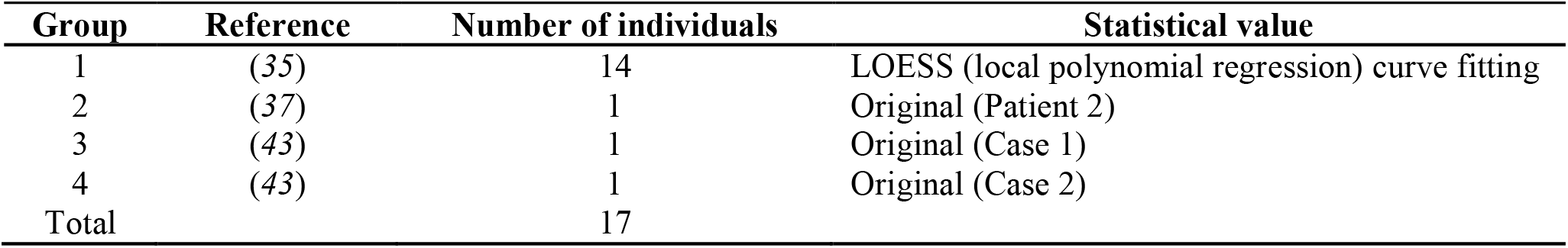
Merged clinical data of CD19^-^ relapse patient groups from individuals.

**Figure S1.**
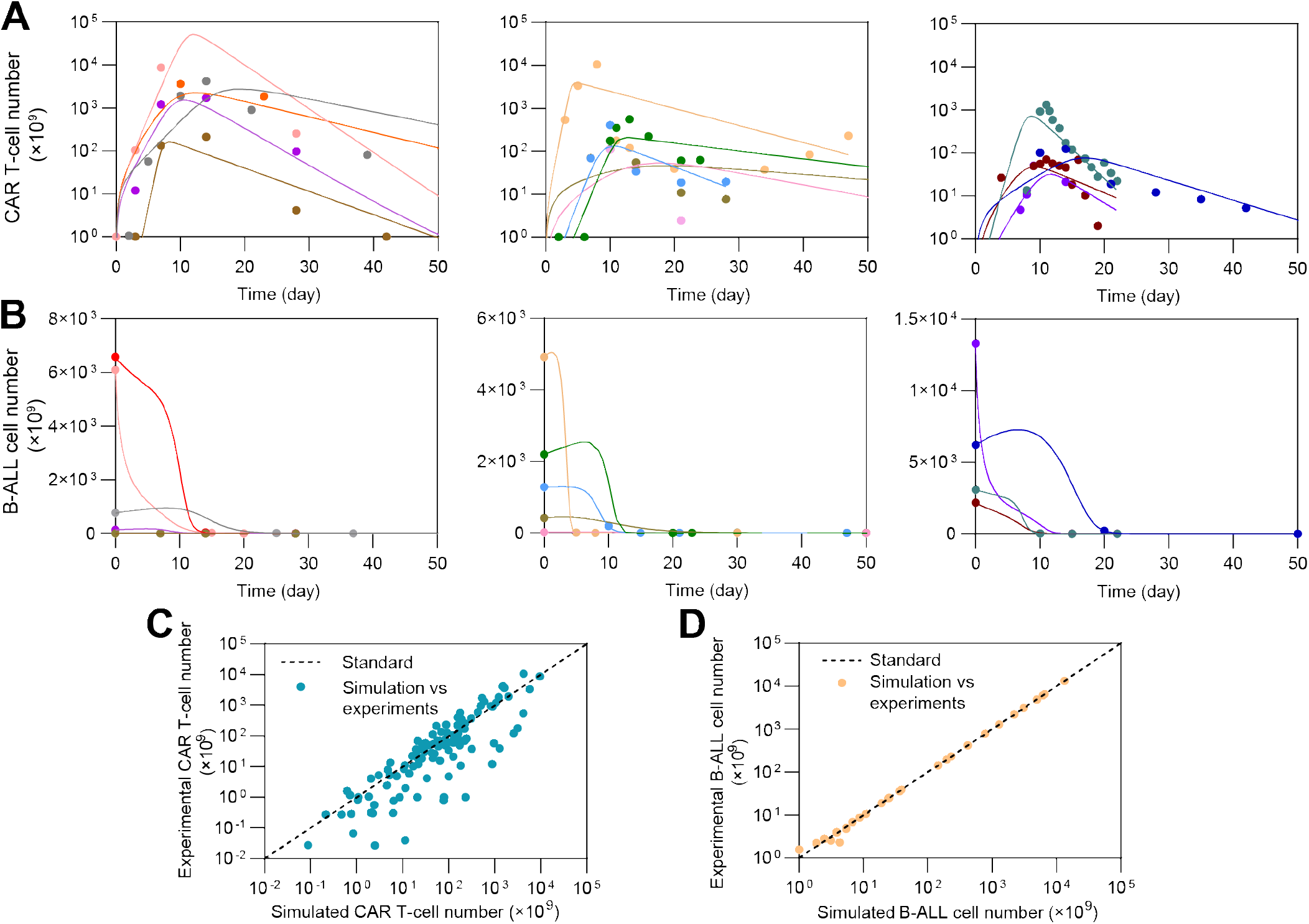
Individual fitting results of continuous remission patients. (**A&B**) Fitting results of CAR T-cell (**A**) and B-ALL cell (**B**) dynamics. The dots represent the experimental data and the lines represent the fitted curves. (**C&D**) Simulated versus experimental results of CAR T-cell (**C**) and B-ALL cell (**D**). Each dot represents the comparison between experimental CAR T-cell number and corresponding simulated result on the same day, and the dash lines represent the standard curves where simulated results are equal to experimental results.

**Figure S2.**
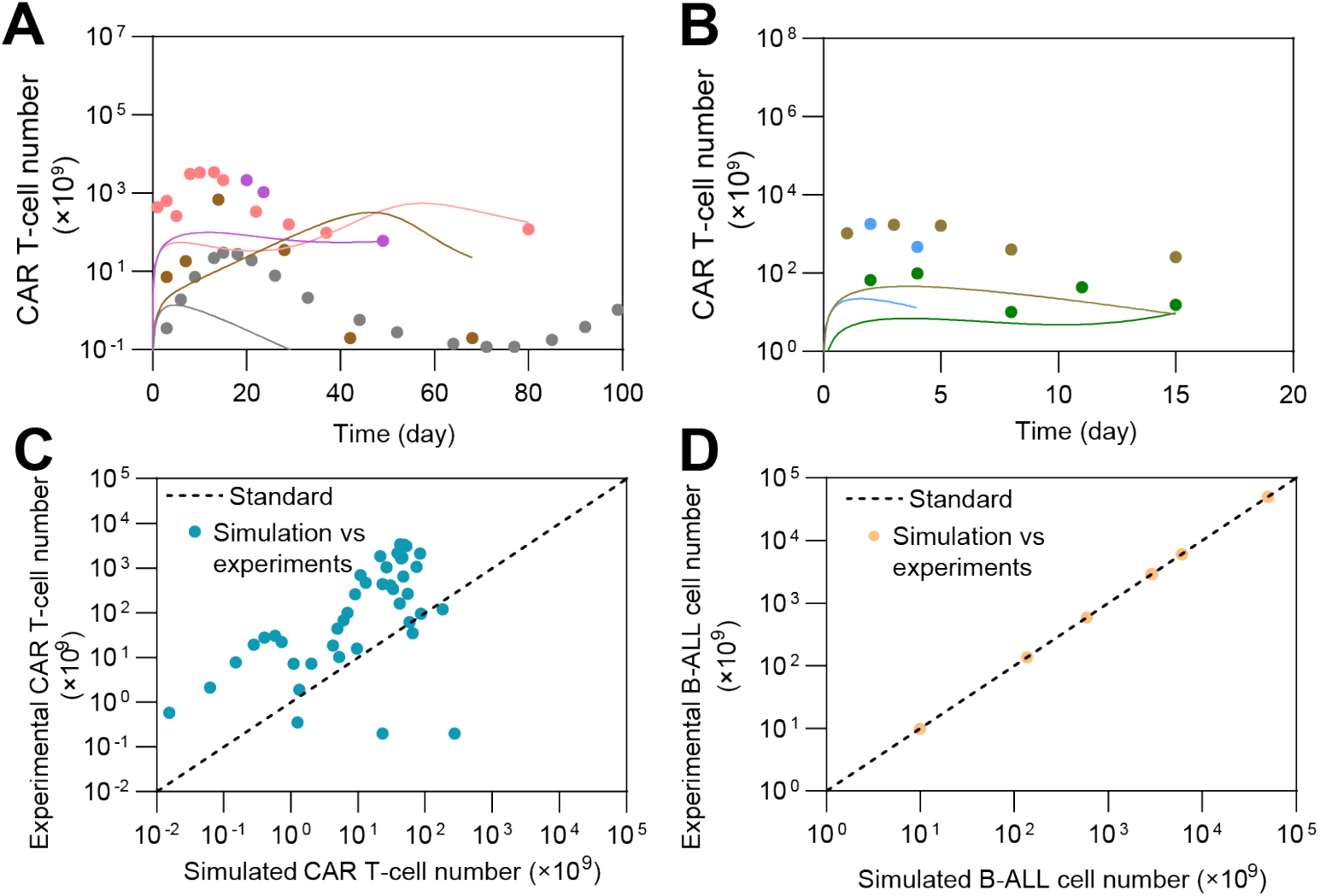
Individual fitting results of non-responding patients. (**A&B**) Fitting results of CAR T-cell dynamics. The dots represent the experimental data and the lines represent the fitted curves. (**C&D**) Simulated versus experimental results of CAR T-cell (**C**) and B-ALL cell (**D**). Each dot represents the comparison between experimental CAR T-cell number and corresponding simulated result on the same day, and the dash lines represent the standard curves where simulated results are equal to experimental results.

**Figure S3.**
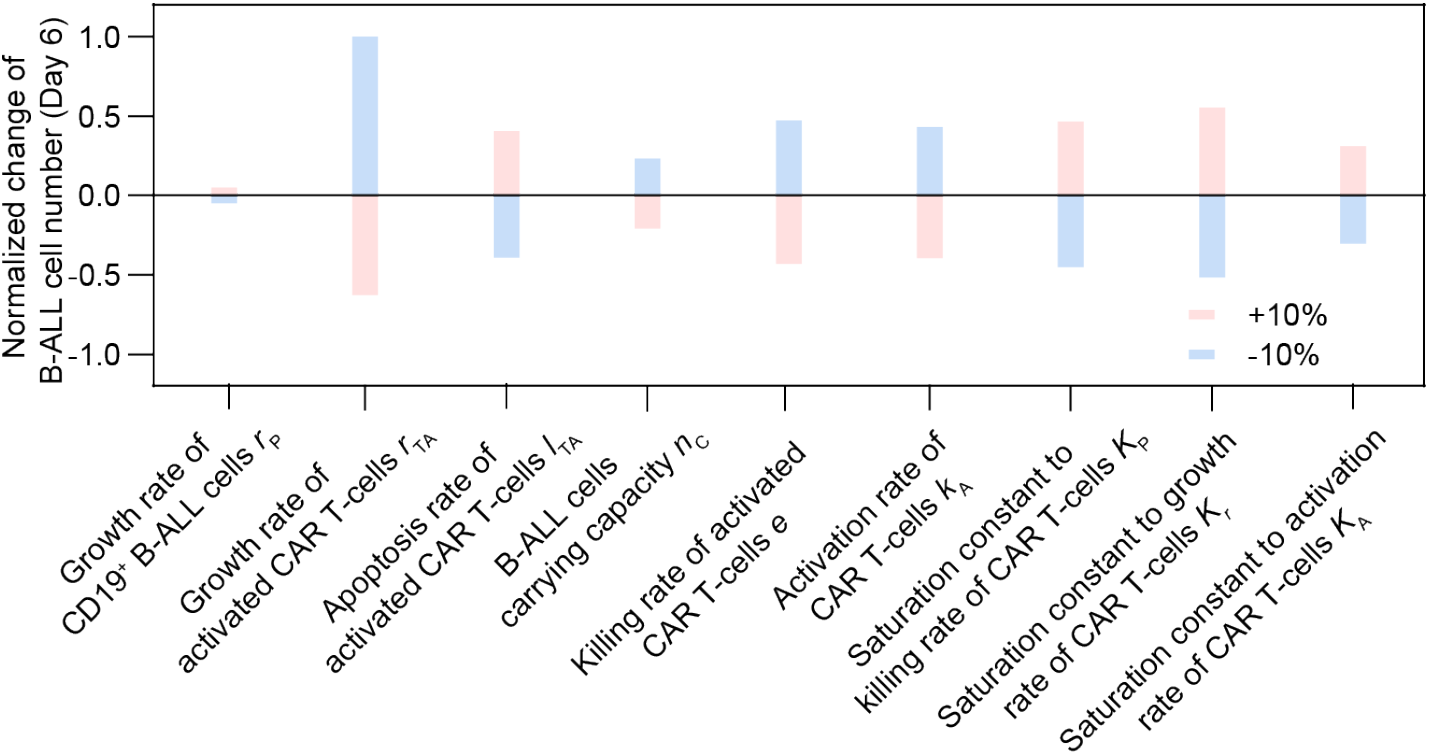
Sensitivity analysis of different CAR T-cell parameters in the computational model.

**Figure S4.**
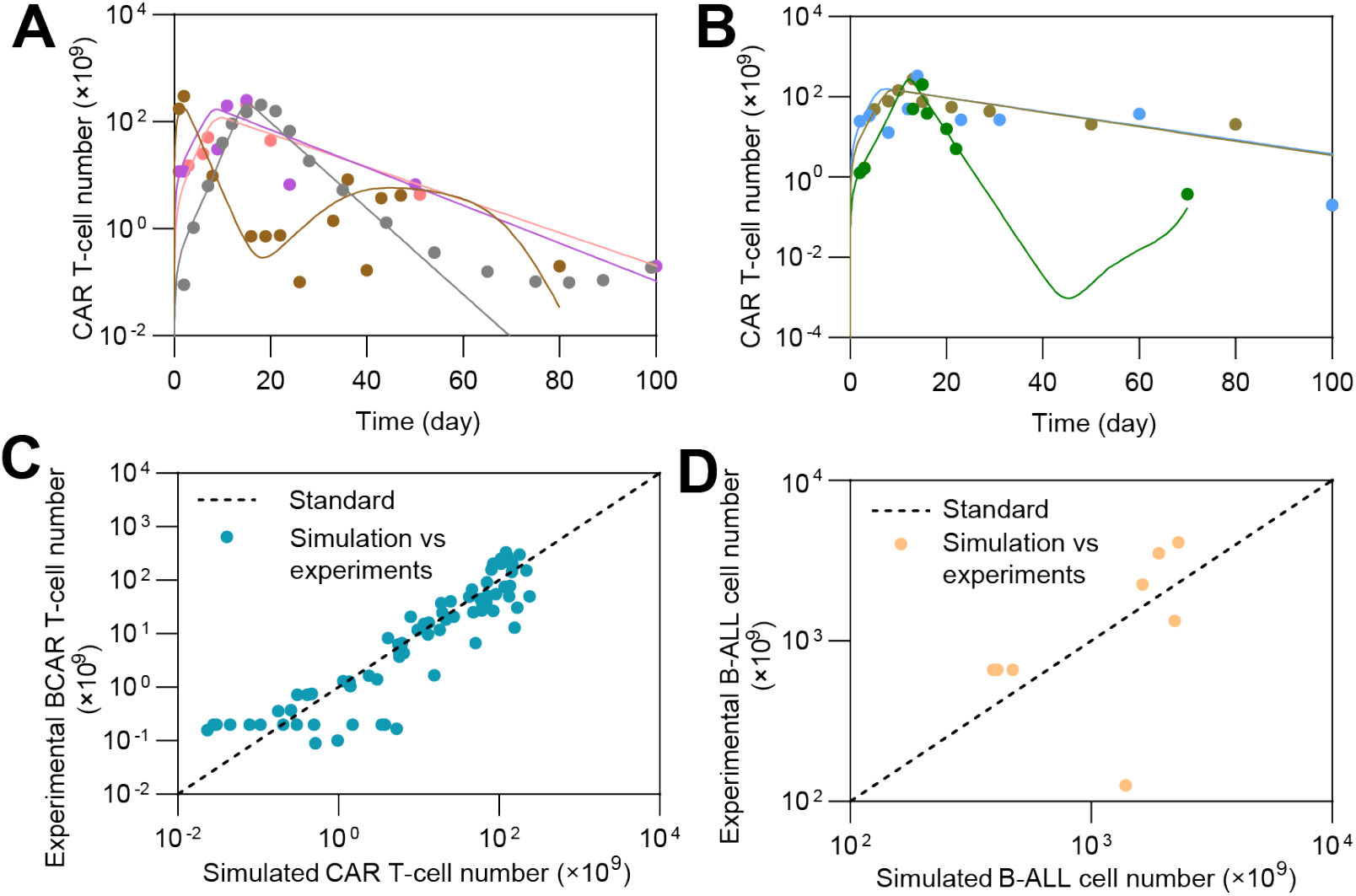
Individual fitting results of CD19^+^ relapse patients. (**A&B**) Fitting results of CAR T-cell dynamics. The dots represent the experimental data and the lines represent the fitted curves. (**C&D**) Simulation versus experimental results of CAR T-cells (**C**) and B-ALL cells (**D**). Each dot represents the comparison between experimental CAR T-cell number and corresponding simulated result on the same day, and the dash lines represent the standard curves where simulated results are equal to experimental results.

**Figure S5.**
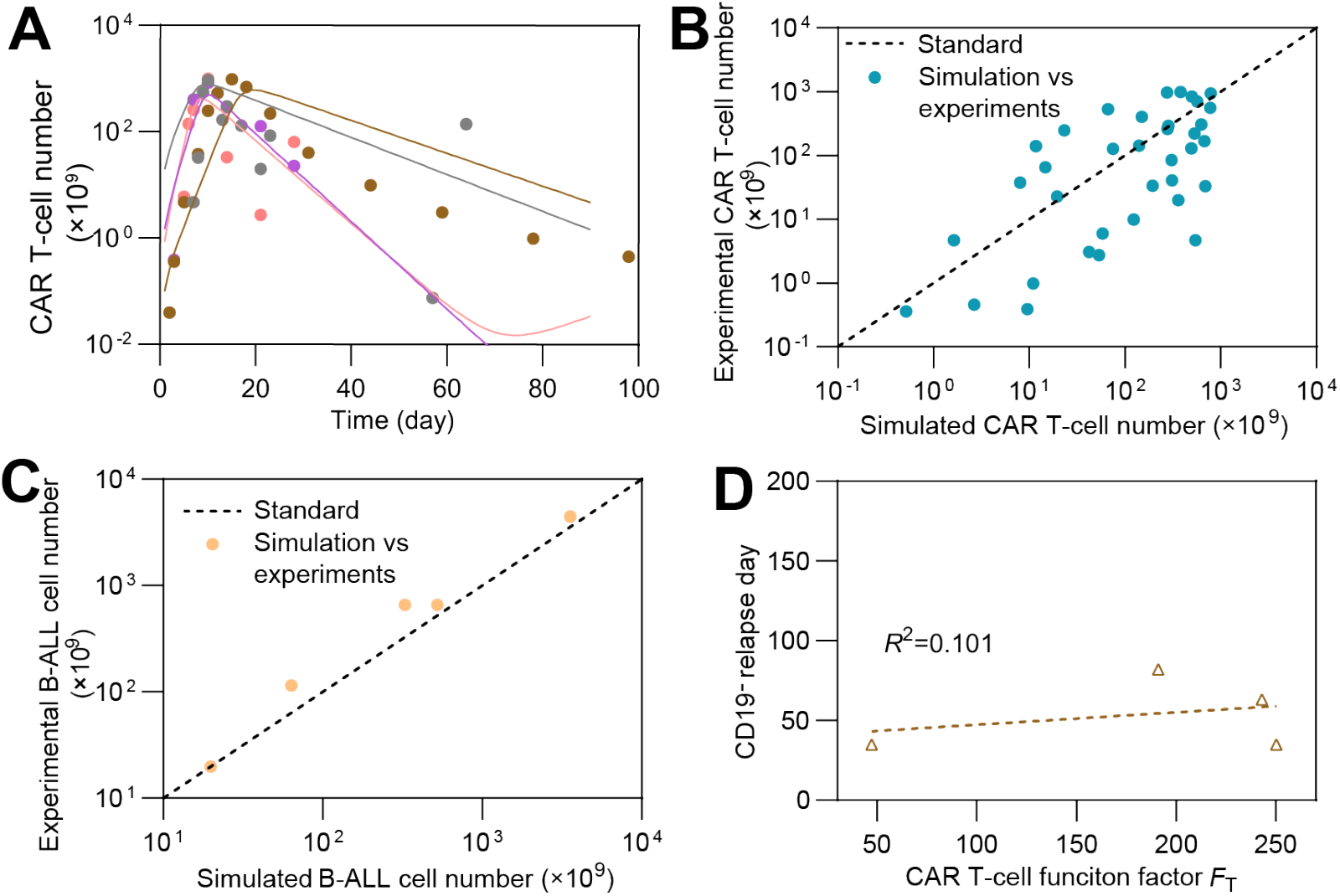
Fitting results of CD19^-^ relapse patients. (**A**) Individual fitting results of CAR T-cell dynamics. The dots represent the experimental data and the lines represent the fitted curves. (**B&C**) Simulation vs. experimental results of CAR T-cell (**B**) and (**C**) B-ALL cell. Each dot represents the comparison between experimental CAR T-cell number and corresponding simulated result on the same day, and the dash lines represent the standard curves where simulated results are equal to experimental results. (**D**) The relationship between CAR T-cell function factor *F*_T_ of clinical patient and CD19^-^ relapse day.

**Figure S6.**
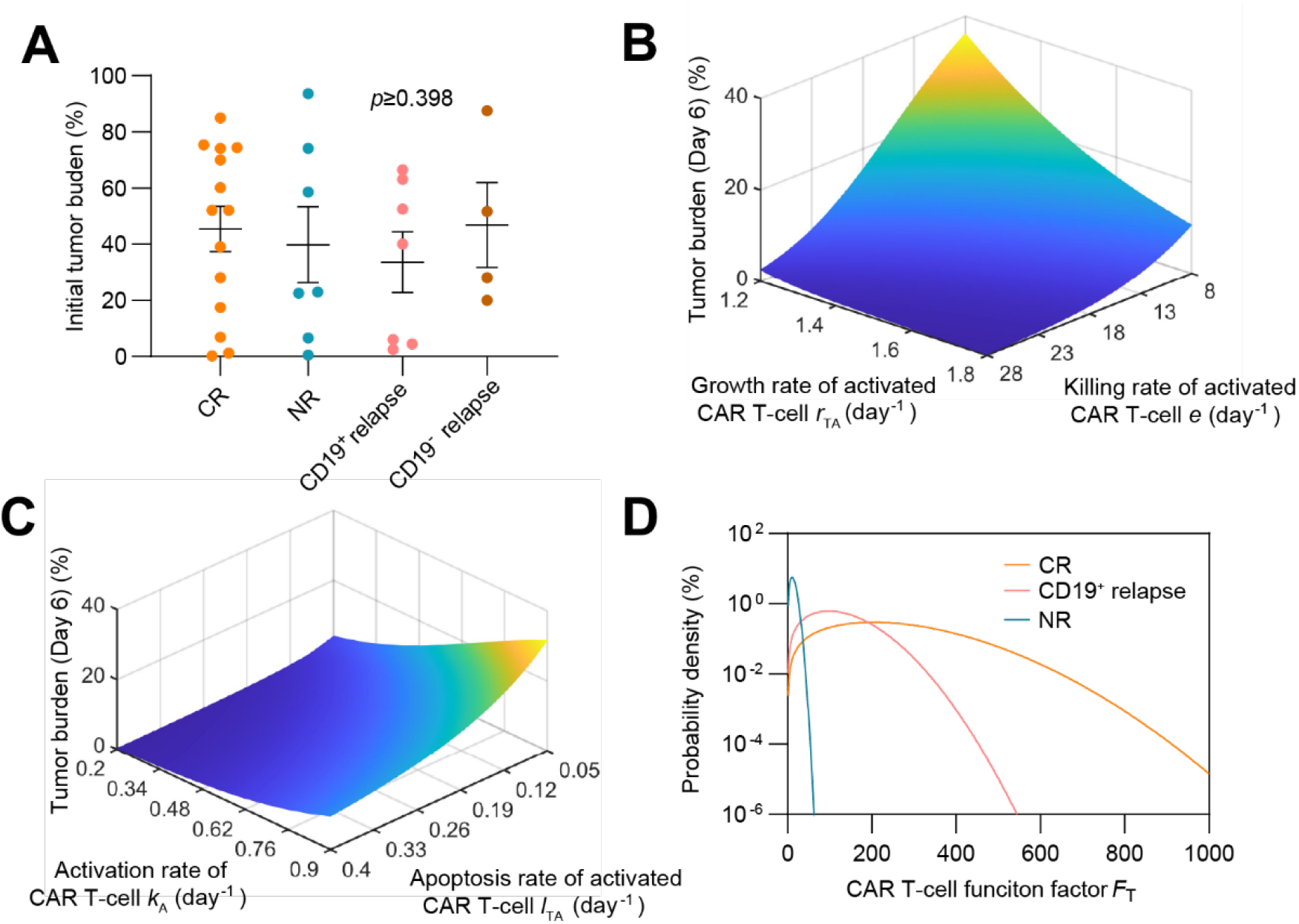
The relationship between CAR T-cell function factor *F*_T_ and CAR T-cell therapy outcome. (**A**) Initial tumor burden in patients of different responses, showing no significant difference (*p*≥0.398). The dots represent the experimental data and the error bars represent the fitted results. Error bars are mean ± SEM. (**B&C**) Effects of *F*_T_ related parameters to therapeutic efficacy: variations of tumor burden at day 6 (day of CAR T-cell at peak) as CAR T-cell growth rate *r*_TA_ and killing rate *e* (**B**), and activation rate *k*_A_ and apoptosis rate *l*_TA_ (**C**) change. The parameters are based on population-level continuous remission patients. (**D**) Probability density of different responses as *F*_T_ changes.

**Figure S7.**
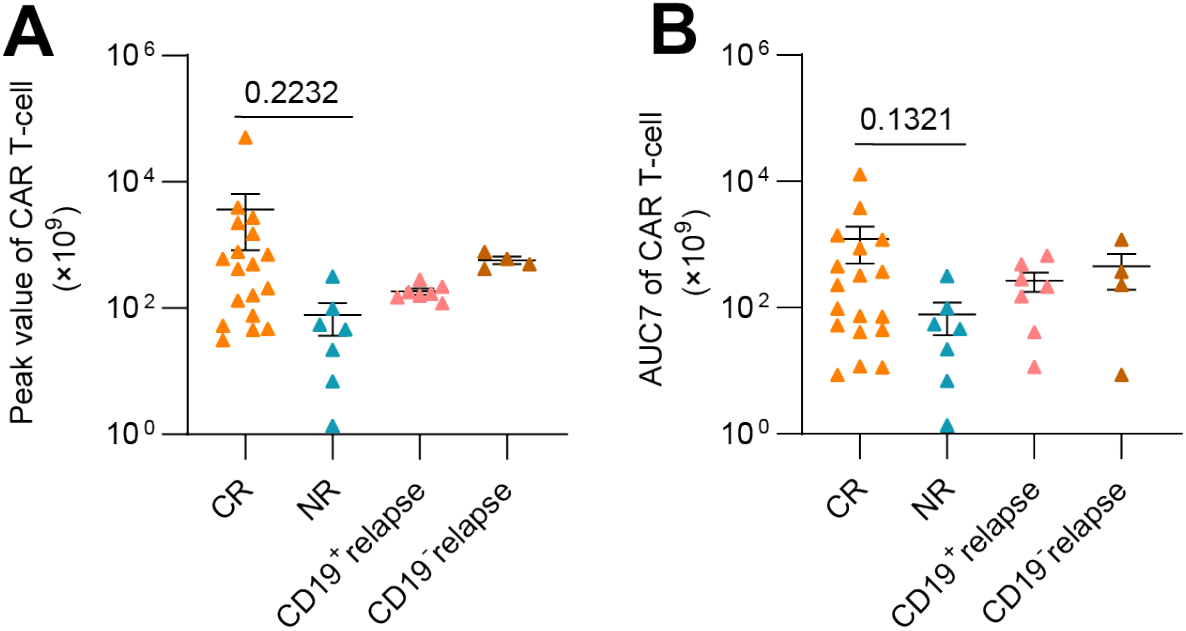
(**A**) Peak value and (**B**) AUC7 of CAR T-cell of different responses to CAR T-cell therapy. Dots and error bars are based on simulated results. Non-significant difference exists between groups for each parameter. Error bars are mean ± SEM. *P*-values were calculated using Welch’s t-test.

**Figure S8.**
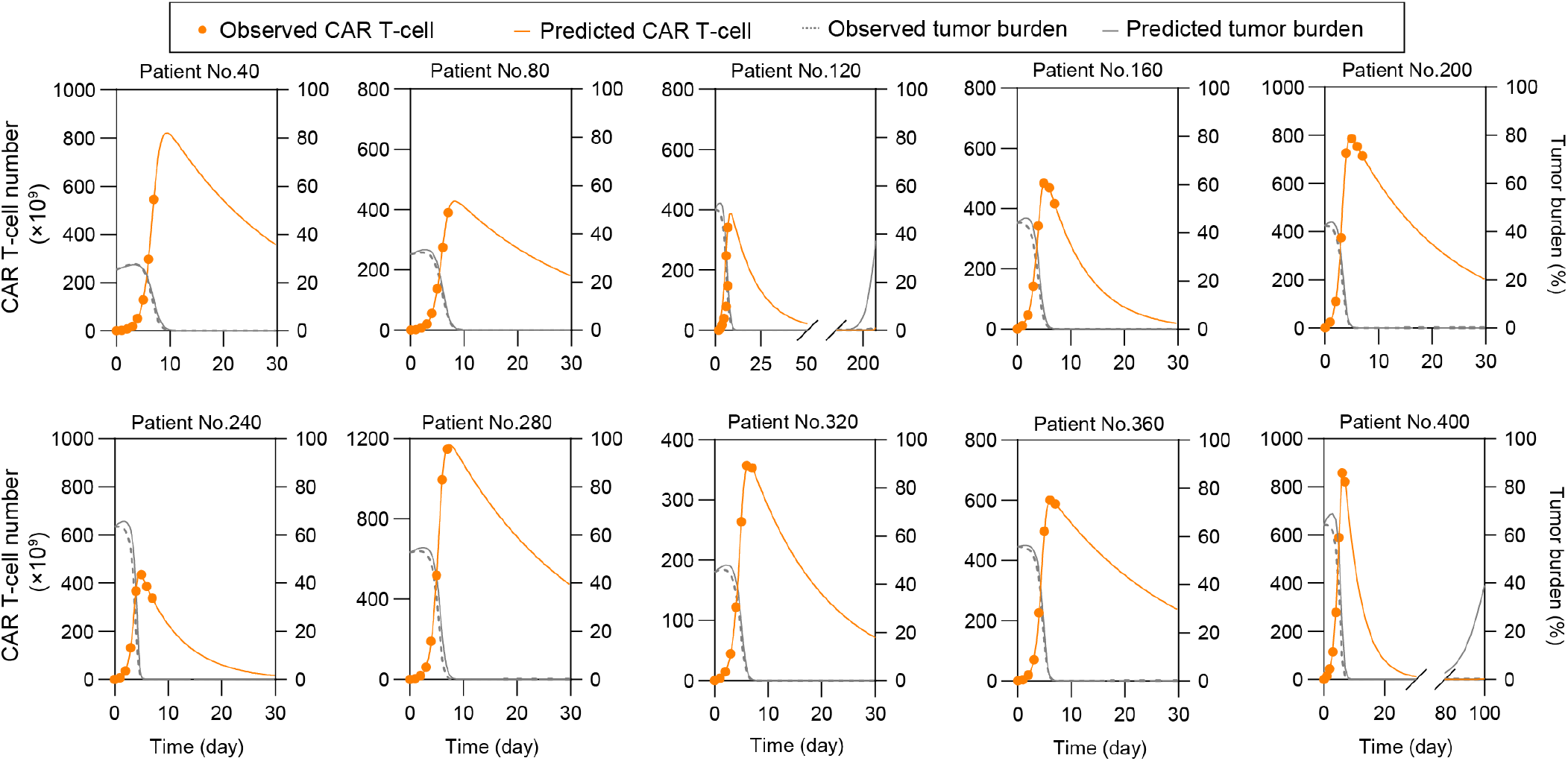
Part of real-time prediction results of CR&CD19^-^ relapse based on virtual patient cohorts. Repetitive virtual patients were chosen at an interval of 40 from the 400 tested patients.

**Figure S9.**
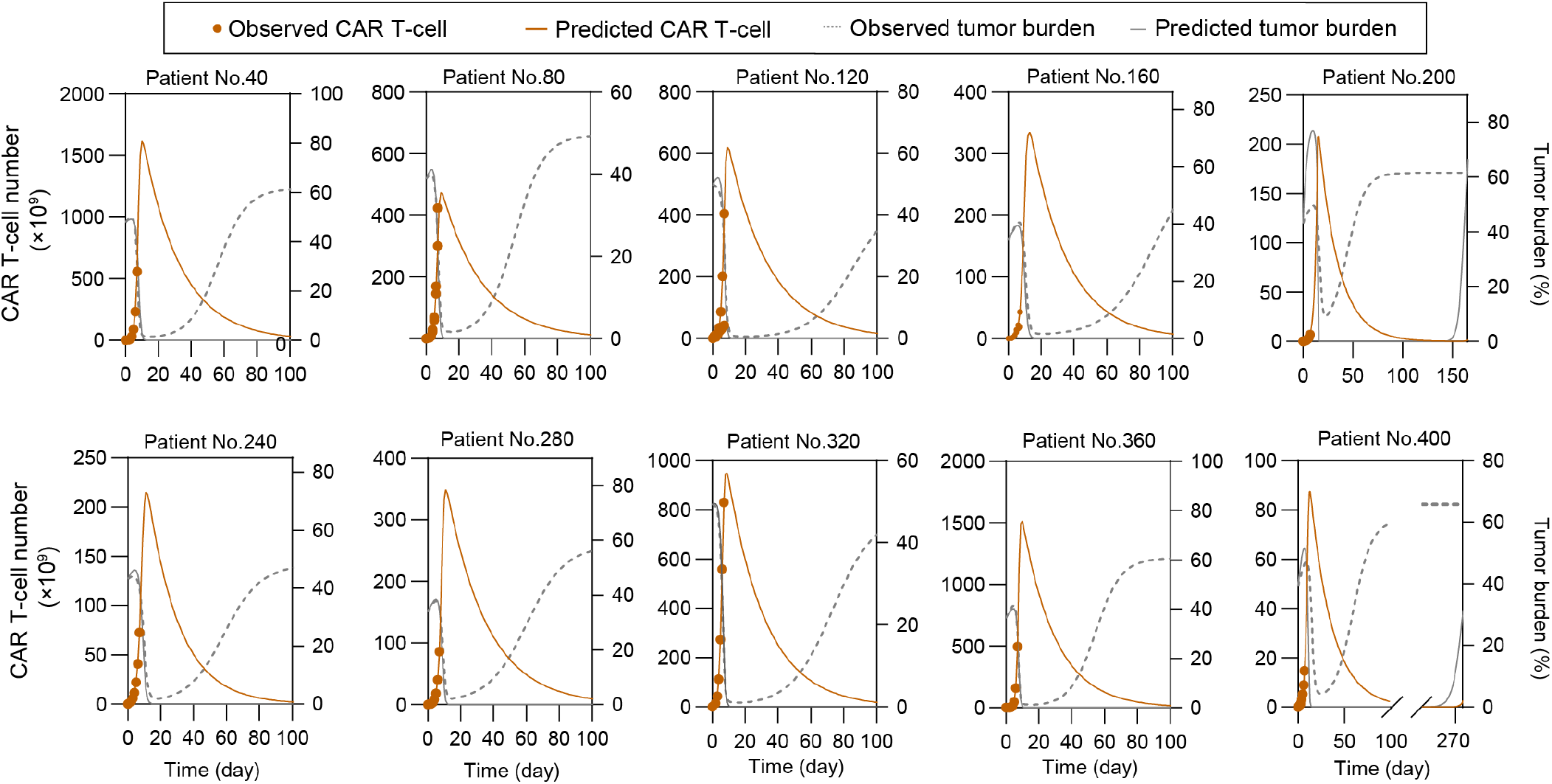
Part of real-time prediction results of CD19^-^ relapse based on virtual patient cohorts. Repetitive virtual patients were chosen at an interval of 40 from the 400 tested patients.

**Figure S10.**
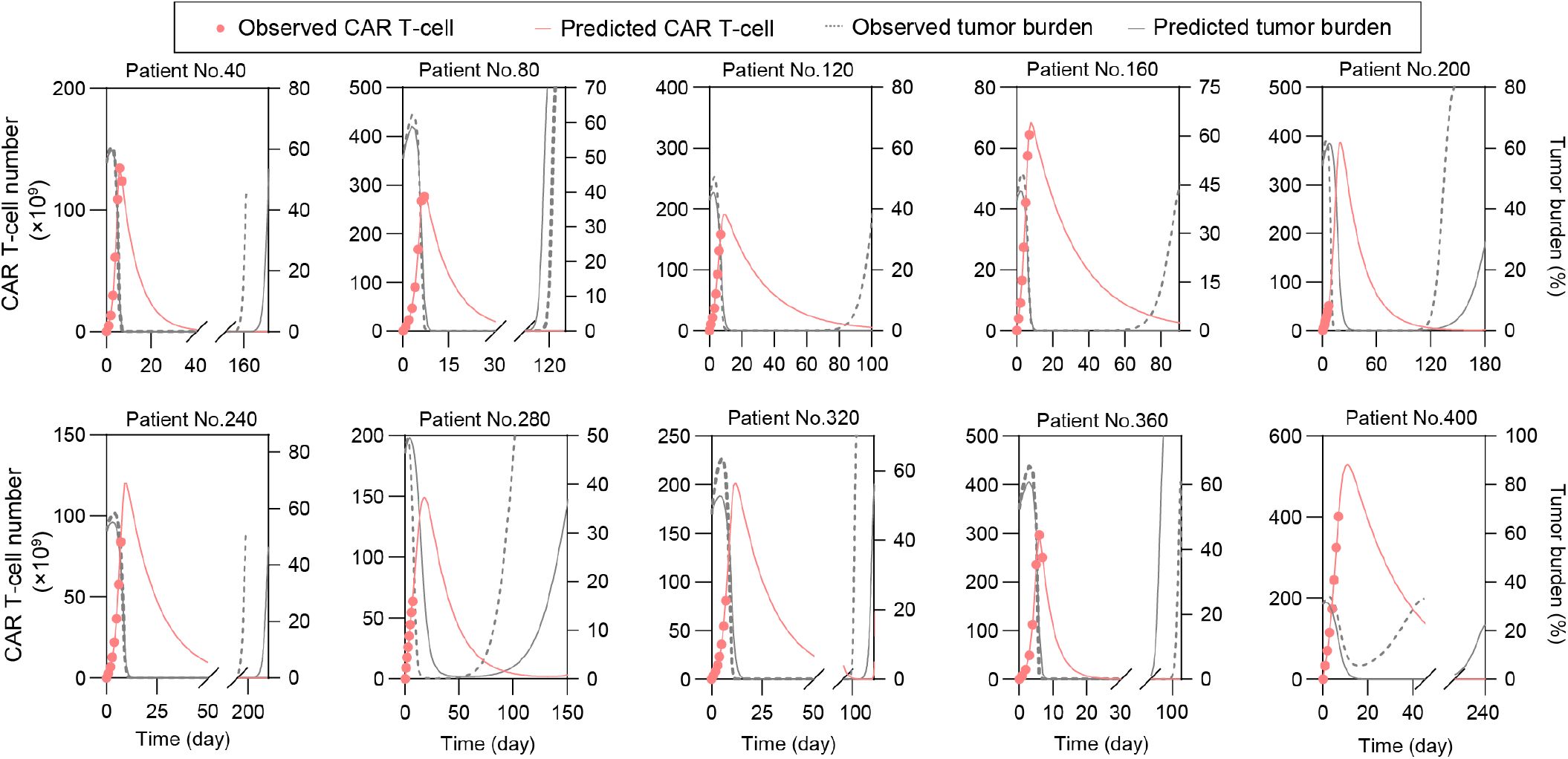
Part of real-time prediction results of CD19^+^ relapse based on virtual patient cohorts. Repetitive virtual patients were chosen at an interval of 40 from the 400 tested patients.

**Figure S11.**
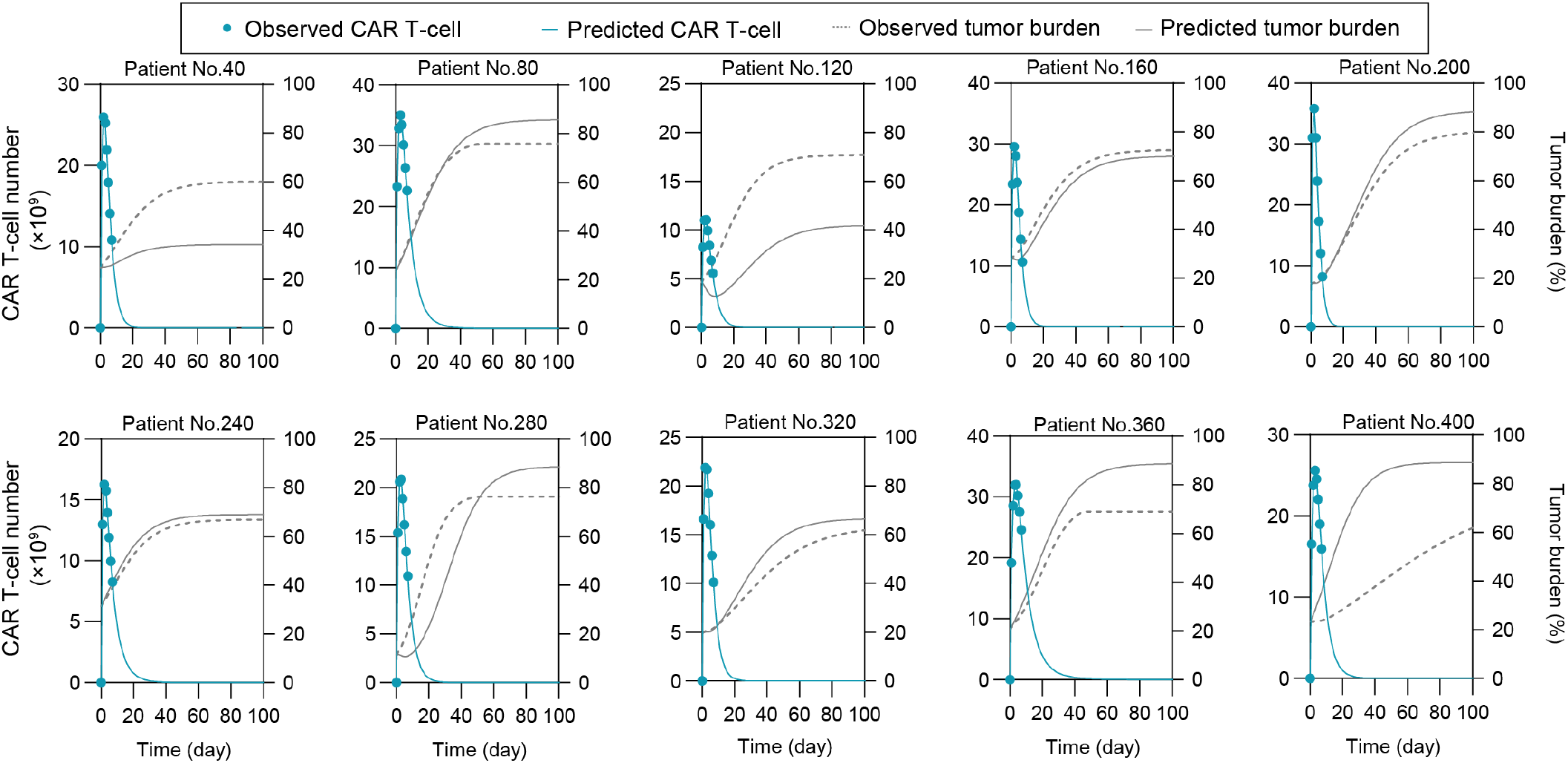
Part of real-time prediction results of non-response based on virtual patient cohorts. Repetitive virtual patients were chosen at an interval of 40 from the 400 tested patients.

**Figure S12.**
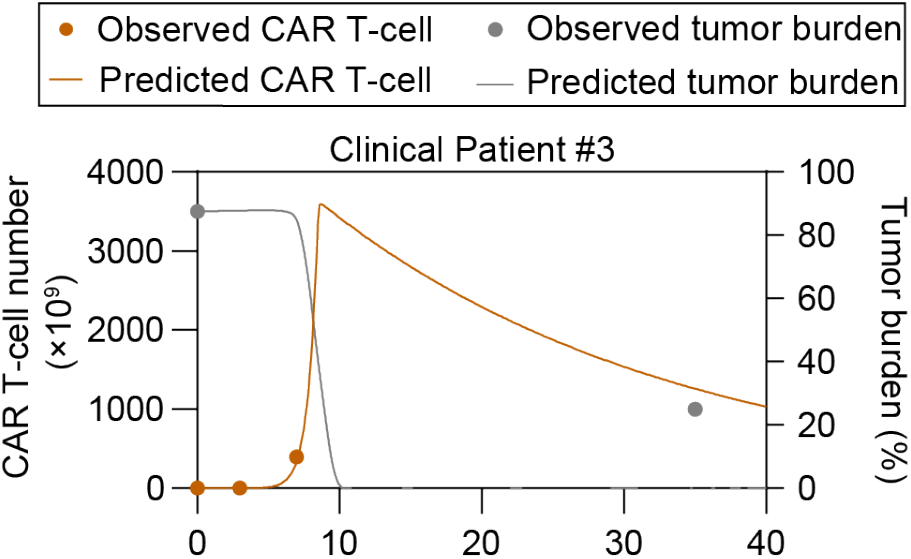
Prediction results based on clinical patient of CD19^-^ relapse.

**Figure S13.**
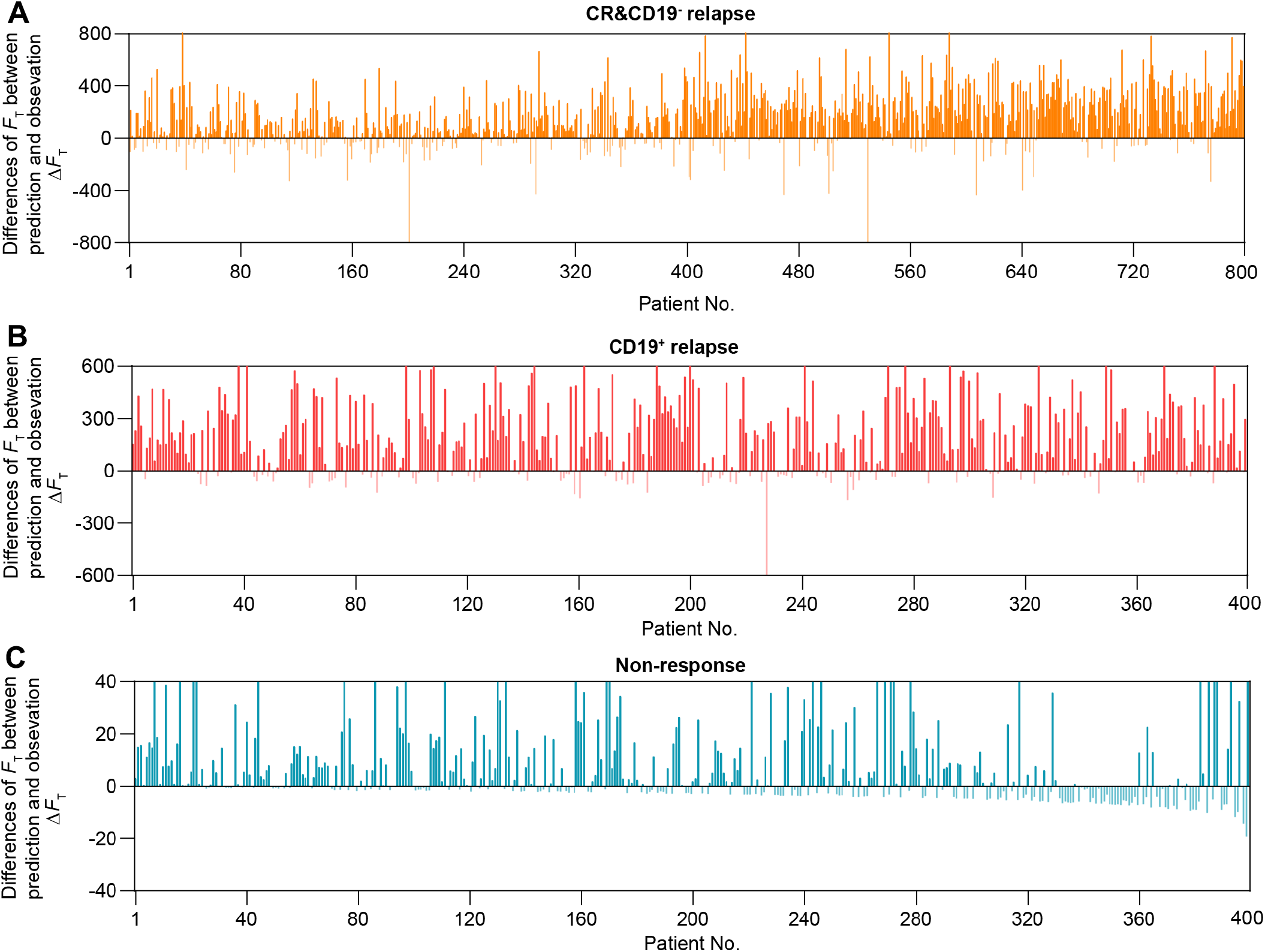
Differences of *F*_T_ between prediction and observation Δ*F*_T_ of CR&CD19^-^ relapse (A), CD19+ relapse (B) and non-response (C) of the 1600 tested virtual patients.

**Figure S14.**
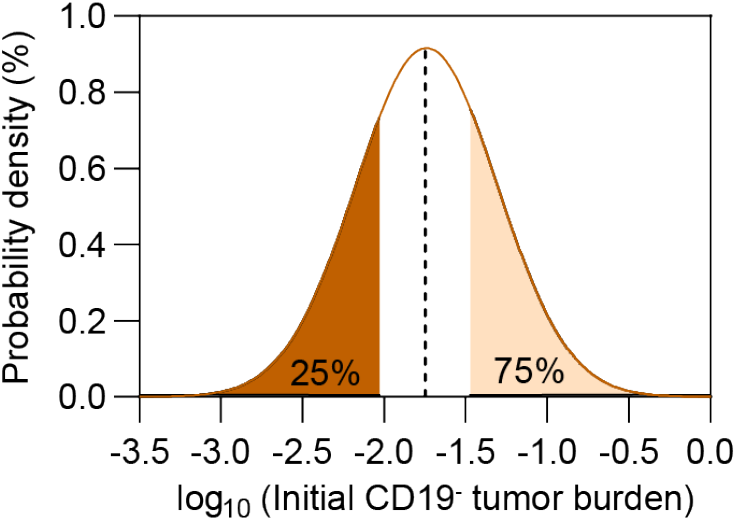
Probability density of the occurrence of CD19-relapse of the 400 tested virtual patients as the value of initial CD19^-^ tumor burden changes.

**Figure S15.**
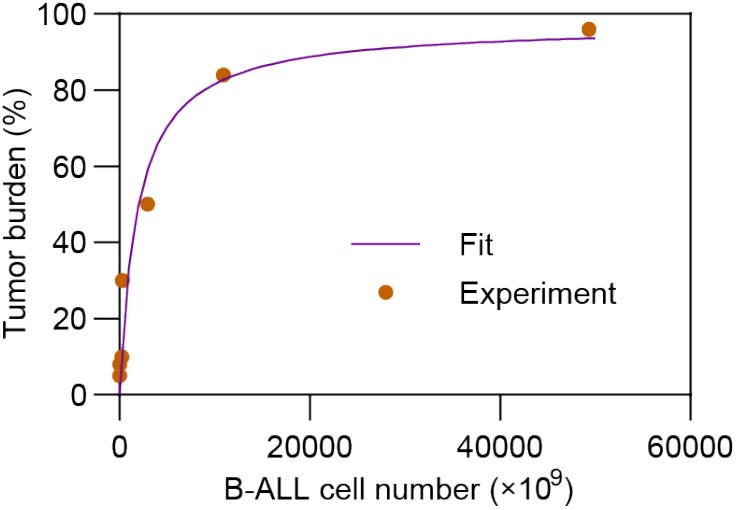
Relationship between the number of B-ALL cell and tumor burden (%). The fitting curve is based on the clinical pre-treatment tumor burdens (**Table 1** of ref. (*25*)) and absolute numbers of B-ALL cells (**Figure 1G** of ref.(*25*)) of MRD^-^ patients.

